# An HERV-W *ENV* transcription in atypical memory B cells linked to COVID-19 evolution and risk for long COVID can express the encoded protein from a ribosome readthrough of mRNA from chromosome X

**DOI:** 10.1101/2024.07.03.24309822

**Authors:** Joanna Brunel, Julien Paganini, Melissa Galloux, Benjamin Charvet, Hervé Perron

## Abstract

The human genome comprises 8% of endogenous retroviruses (HERVs). Though HERVS contribute to physiological functions, copies retained pathogenic potential. The HERV-W ENV protein was shown expressed in patients with worse COVID-19 symptoms and post-COVID syndrome. A significant detection of the mRNA encoding HERV-W ENV from patients with COVID-19 in B cells from RNAseq reads obtained from peripheral blond mononuclear cells. This data stratified with increased COVID-19 symptoms or with post-acute sequelae of COVID-19 (long COVID) after 3 months. The HERV-W *ENV-U3R* RNA was confirmed to display the best alignment with chromosome X ERVWE2 locus. However, a stop codon precluding its translation was re-addressed after recent understandings of ribosome readthrough mechanisms. Experimental results evidenced that this HERV gene can effectively express a full-length protein in the presence of molecules allowing translation via a readthrough mechanism at the ribosome level. Results not only confirm HERV-W *ENV* RNA origin in these patients, but show for the first time how a defective HERV copy can be translated into a complete protein when specific factors make it possible at the ribosome level. The present proof of concept now requires further studies to identify the factors involved in this newly understood mechanism, following SARS-CoV-2 exposure.

## 1. Introduction

The COVID-19 pandemic has created what could be described as a secondary pandemic, with the onset and persistence of many symptoms after the viral infection is resolved, affecting about 10-20% of patients after COVID-19 ^1–3^. This is now a major concern for millions of patients around the world, and many of them evolve toward chronicity^4^. This post-infectious syndrome has been referred to as post-acute sequelae of COVID-19 (PASC), post-COVID syndrome/condition or, more commonly now, long COVID (LC)^5^. The appearance of so many cases within a short delay after COVID-19, the frequency, and the important number of symptoms revealed the involvement of a viral infection in the appearance of a post-infectious syndrome much different from that the fatigue of a convalescence phase^6,7^.

Beyond, it questions the etiopathogenesis of chronic pathologies whose initial or underlying cause remains unknown and whose epidemiology tells us that they may be associated with infections contracted before their onset^8–11^. However, unlike COVID-19, it seems that the delay between the activating role of an infectious agent and the appearance of major or detectable clinical symptoms can be much longer than for the onset of post-COVID symptoms. This probably explains the difficulty of linking certain infections with post-infectious pathologies observed later when the agent that induced their development in susceptible individuals (linked to genetics, comorbidity or exposure to certain environmental factors), is no longer or rarely present^12–15^.

In this context, studies that we have conducted for many years, along with those of other groups, have shown that infectious agents can dysregulate the epigenetic control and, also, activate the abnormal expression of human endogenous retroviral elements -HERVs-^16–51^. HERVs constitute 8% of the human genome^52^ and, altogether with transposable or retro-transposable (TE) elements they represent about a half of the human DNA, now referred to as the “dark genome”^53–55^. This must be compared to the small proportion (1-3%) of sequences encoding somatic and physiological proteins of the human body ^56–58^.

The high number of highly homologous copies of the different families of TEs and HERVs inserted into human DNA, as well as their copy-number variations in human individuals^23,59–63^, create a complexity that often exceeds the possibilities of an analysis of precisely localized copies in one or more chromosomal sites. This nonetheless remains possible at the level of families involved in their activation by an infectious agent or in pathologies evolving over the long term. Furthermore, when a HERV protein is abnormally produced and causes pathogenic effects, its related HERV family can also be detected and characterized ^18,20,21,64^.

This is why, after it had been shown that the retroviral envelope protein of the HERV-W (W-ENV) family had pathogenic effects on immune and neurobiological functions^21,24,31,39,65,66^, but also that HERV-W ENV genes could be abnormally activated by viruses or parasites inducing its long-lasting expression^25,35,38,40^, the question of its deleterious expression was raised in the context of COVID-19. In the absence of prior neutralizing immunity, COVID-19 was characterized by an hyperactivation of innate immunity causing a “cytokine storm”, in parallel with lymphocyte deficiency and impairment of adaptive immunity functions that are essential for infection control^67^. The W-ENV protein was previously called MSRV-ENV after its discovery in multiple sclerosis, and then pHERV-W ENV for its role in its pathogenesis and that of other diseases ^22,27,43,68^. W-ENV is a potent agonist of the TLR4 receptor ^39,69^ and, therefore, of the pro-inflammatory activation of innate immunity. In addition, and although this is conditional on the prior activation of TLR4 signaling pathways, this protein showed superantigen-like activity on T cells, in which it had never been observed to be expressed ^65^.

In a first study carried out during the first wave of the COVID-19 pandemic, the transcriptional activation and HERV-W ENV protein expression was demonstrated from the onset of infection in peripheral blood lymphoid cells and, for the first time, in T cells^70^. With a retrospective follow-up of the patients studied, this study showed that a high percentage of lymphocytes expressing W-ENV predicted a severe evolution of COVID-19 with cytokine storm and a need for oxygen or even intubation in intensive care. A second study ^17^ subsequently showed that: (i) *in vitro* exposure of healthy control peripheral blood mononuclear cells (PBMC) to SARS-CoV-2 could induce transcriptional activation and then protein expression of W-ENV in about 20% of PBMC from normal blood donors, (ii) in early cohorts collected during the PCR diagnosis of COVID-19 the soluble W-ENV protein was found in the same proportion (about 20%) of sera from patients, but their further clinical evolution was not known (iii) that the proportion of patients with soluble W-ENV in the serum increased to 100% in patients hospitalized for severe forms in the intensive care unit (ICU) during successive epidemic waves with different variants and that (iv) autopsy tissues of COVID-19 patients who died of severe forms showed high W-ENV expression in cells close to, but different from, those that expressed SARS-CoV-2 antigens; namely: alveolar macrophages and pulmonary subepithelial lymphoid infiltrates; endothelial cells of certain blood vessels in all analyzed organs and the only positive cells in cardiac and peri-cardiac tissues; cells associated with intra-vascular blood clots; lymphoid infiltrates of the epithelial tissues and villi of the digestive tract and, massively, in lymphoid structures associated with mucous membranes; finally, microglial cells of the cerebral parenchyma with a complete absence of SARS-CoV-2 antigen in the central nervous tissue whereas, in sections at the level of the cribriform plate separating the olfactory bulb from the nasal mucosa, SARS-CoV-2 proteins were strongly detected in neighboring mucosal cells as well as W-ENV in lymphoid infiltrates.

Subsequently, the discovery of a post-COVID (PASC) or long COVID (LC) syndrome raised the question of a possibly persistent W-ENV expression following its initial activation. Its known effects involving the persistent activation of cerebral microglia and various immune dysfunctions, was therefore calling for further research in LC. Initial results showed that about one-third of patients with LC had a detectable W-ENV protein in their serum and, therefore, that the expression of this protein persisted at least in certain tissues with soluble W-ENV secretion found in peripheral blood ^71–73^, even if no longer in circulating lymphocyte cells as observed during the acute COVID-19 infection. It is therefore necessary to detect and confirm the abnormal activation of this expression and, if possible, from the early SARS-CoV-2 infection phase. It should anticipate W-ENV expression that has already been shown to be pejorative for the course and complications of COVID-19 itself ^70^ and to be involved in a significant proportion of patients who developed LC associating a persistent expression of this pathogenic W-ENV protein ^72,73^.

To firstly address this pathogenic expression, we were interested in studies carried out on the PRESCO cohort of COVID-19 patients whose blood samples were collected early during SARS-CoV-2 infection and who were clinically followed up to the post-infectious phase with or without an LC diagnosis at this stage ^74^. While immunological and multi-omics analyses showed different profiles without effective specifications for targeted therapeutic approaches^75,76^, the clinical follow-up of a large subgroup of patients made it possible to identify three clusters with three divergent evolution of symptoms over a follow-up of several months^74^: a group (cluster 1) whose symptoms regressed, with spontaneous improvement, a group (cluster 2) whose symptoms persisted but remained stable in number and intensity and a group (cluster 3) whose symptoms worsened with the appearance of new symptoms not observed during the previous visits. This favorable, stable or pejorative evolution has grouped together symptoms related to complications in the infectious phase but also to their number and severity in patients with long COVID.

In the present study, we first analyzed the sequences extracted from samples with available RNAseq data of patients who had a clinical follow-up up to the last post-infection visit (visit 5), to confirm the analyzable coverage of the sequences of interest and to compare their expression levels in all the cell phenotypes studied. This was compared between patients classified in the three clinical clusters as well as in the patients diagnosed as LC (PASC) or non-LC (no-PASC) at the end of this follow-up. The pilot study on a few cases led us to analyze the large amount of data available from RNA sequencing during visit 1 and 2 (early infection) of all patients followed clinically and classified in cluster 3 of pejorative course in visit 5, versus cluster 1 of favorable spontaneous course. We also analyzed the complications (reported symptoms) corresponding to the COVID-19 infection as considered within clinical clusters, separately from the post-infectious status with or without LC.

Then having confirmed the mRNA sequence associated with this pathogenic expression of HERV-W *ENV*, we re-addressed the question of the chromosomal copy at the origin of this RNA transcription. As previously suggested, the best homology over the entire sequence was supposed to be located on chromosome Xq22.3, but the corresponding open reading frame (orf) was confirmed to have a premature stop codon that could only produce an N-terminally truncated protein^77^, which was not compatible with the molecular weight and biochemical characteristics of the HERV-W ENV protein analyzed from MS brain lesions^20^ and the labelling with antibodies targeting distant epitopes of its amino-acid sequence ^34^. This locus, now named ERVWE2, was also confirmed to retain this stop codon in all healthy and MS individuals in a dedicated study on human DNA^78^. This led to debate on hypotheses consisting in a transcriptional interference between copies from multiple chromosomes, in the presence of non-ubiquitous copy(ies) in genetically susceptible individuals, or in the induction of somatic DNA rearrangements allowing the complete HERV-W protein(s) translation ^79,80^.

In this study, we have shown for the first time that the RNA corresponding to this ERVWE2 sequence can be translated into a full-length HERV-W ENV protein as described in pathogenic conditions, provided that specific helper molecules interact with the ribosomal machinery to allow a readthrough translation replacing the stop codon by a tryptophan. How one HERV-W copy may encode this pathogenic ENV protein is experimentally shown. This new understanding of an unusual human gene transcription is analyzed from the structure of the RBM41 gene intron and exon across which ERVWE2 copy is inserted.

## 2. Materials and methods

To conduct a study on the PRESCO cohort, were retrieved RNA sequencing data from the blood mononuclear cell subpopulations (PBMCs) of patients that were sorted and sequenced with the Verily’s Immune Profiler platform ^76^. Our analyses focused on all the sequences of the HERV-W genome, including those of the gene encoding the W-ENV protein (*ENV* according to the nomenclature of human genes, or *ENV*, according to the virological nomenclature), with the expected structure of its mRNA which includes the U3R part of the flanking repeat region (LTR) where the polyadenylation signal is located (**Fig. 1**).

**Figure 1:**
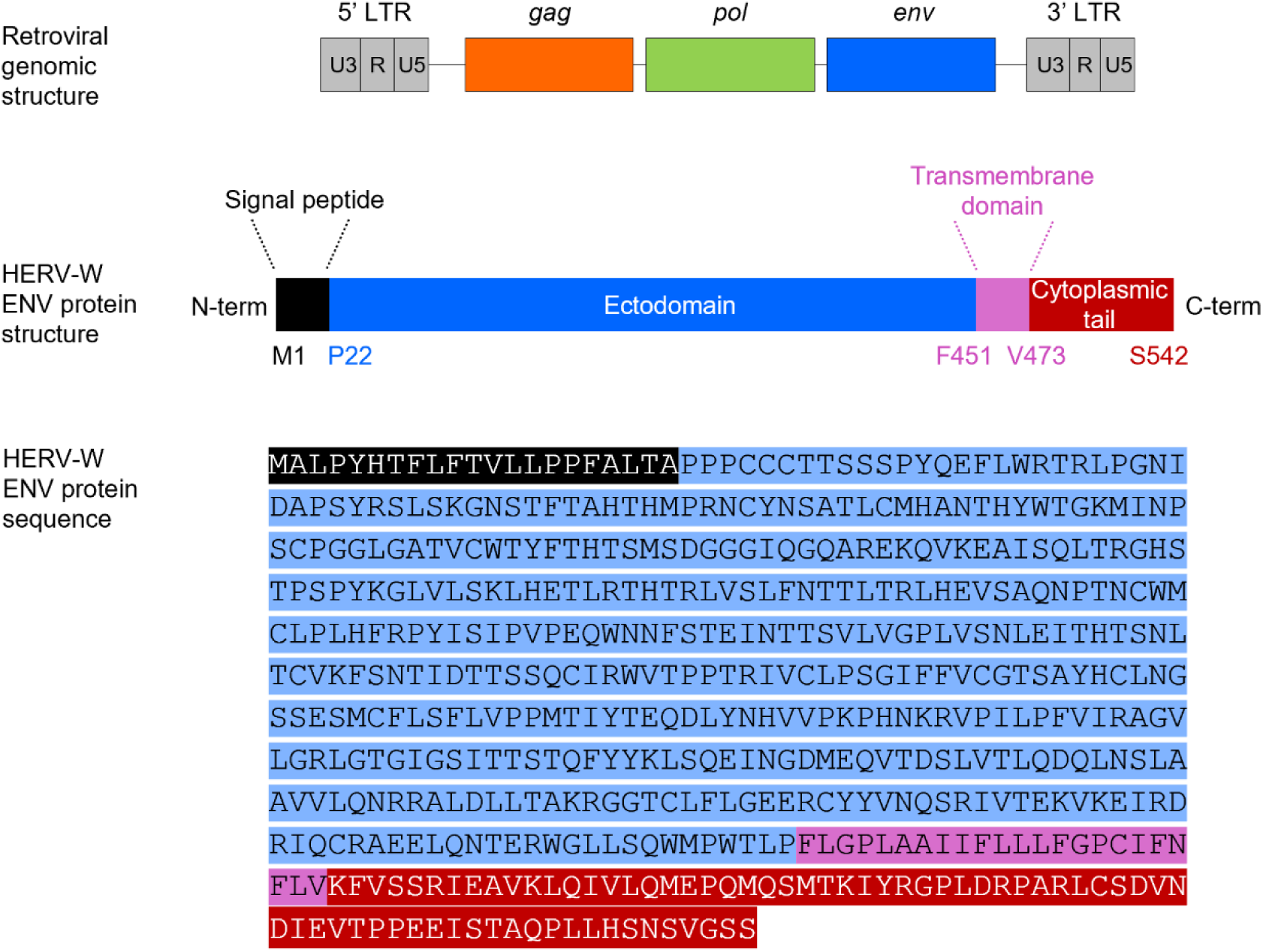
HERV-W Nucleotide structure and envelope protein: **Top:** The nucleotide structure of a retroviral genomic RNA obtained from HERV-W (previously named MSRV) original sequences isolated from multiple sclerosis^68^ comprises long terminal repeats (LTR,; RU5 region in 5’ and U3R region with polyadenylation signal for mRNAs in 3’) flanking the sequences of the gag gene-encoded proteins (Matrix, capsid and nucleocapsid), the pol gene-encoded enzymes (protease, reverse-transcriptase and integrase) followed by the *ENV* gene-encoded envelope protein. **Middle:** The different domains of the envelope protein are represented: signal peptide (propeptide in black), ectodomain in blue, the transmembrane domain encompassing the membrane-anchoring sequences in pink and the cytoplasmic tail in dark red. **Bottom:** The amino acid sequence of HERV-W ENV protein (GenBank: AAK18189.1).

### 2.1 RNAseq data and analyses

#### 2.1.1 Presco project data

The study used the RNA sequencing data of the Presco Project (clinicaltrials.gov NCT04388813) on samples analyzed with Verily’s Immune Profiler platform, which also included RNA sequencing of samples^74–76^. Briefly, this platform begins with the isolation of 25 immune cell subsets (5 myeloid cell subsets, 7 B cell subsets, 10 T cell subsets, 2 Natural Killer cell subsets, and a bulk peripheral blood mononuclear cells sample) from a starting material of approximately 10 million cryo-preserved PBMCs per individual. RNA-seq is performed for each of the 25 subsets. RNA-seq was performed using a SMART-Seq2-based procedure optimized for use on hundreds of cells.

We decided to analyze two subsets of data according to the associated meta information. The composition of datasets is provided below:

#### 2.1.2 RNAseq data from the PRESCO cohort

Data are made available upon requests sent at.

#### 2.1.3 Reference sequence dataset

A reference data of HERV-W sequences of interest was built at fasta format. Sequences were retrieved from NCBI. A fasta file was created for each and a global fasta file including all sequence was also created. List of sequence, details and accession are provided in Supplementary Files.

#### 2.1.4 Reads Extraction

Reads extraction from the PRESCO cohort^76^ was done in two steps. Firstly, a non-stringent analysis was done do retrieve all potentials reads of interest. This analysis was performed using bowtie2 software^81^. This step was done using a local mode alignment. The fasta file containing all HERV-W sequences was used as reference. An example of command line used is provided in Supplementary files. The second step retrieved reads associated with each individual reference, using a more stringent analysis with end-to-end alignment. The same bowtie2 software was used but with an end-to-end alignment mode and, as reference, a fasta file with the sequence of interest only. Input reads used for this analysis correspond to the reads obtained during the first step.

#### 2.1.5 Read quantification

For each file of reads obtained after extraction analysis, we used the fastqc software^82,83^ and the multiQC software^84^ to look at the reads features and to obtain the number of reads per file. We automatically retrieved this information using a simple algorithm developed internally to summarize all results in a table as input data for statistical analyses.

### 2.2 Statistical analyses

#### 2.2.1 PASC and no-PASC comparison for all samples

For each gene, the frequency of reads was expressed as the number of reads for a specific HERV-W gene per one million of total RNAseq reads of the sorted cells^76^ from each sample. To investigate the difference in read frequency, a statistical analysis was conducted using a two-tailed independent samples t-test to compare mean read frequency between PASC and no-PASC patients for each gene and each cell phenotype. Analysis was done using R version 4.2.2 (A Language and Environment for Statistical Computing. R Foundation for Statistical Computing, Vienna, Austria. URL: https://www.R-project.org/) and plots were generated using ggplot2 version 3.4.4 ^85^ and ggpubr version 0.6.0 packages^86^.

#### 2.2.2 Difference in ratio of read frequency for *ENV* gene in aMBc cells between clusters

The ratios of Env reads frequency between aMBc cells and the mean frequency of all other PBMC phenotypes^75^ were computed for each sample from patients previously identified to belong to a clinical cluster with favorable (cluster 1) or pejorative (cluster 3) symptoms evolution after COVID-19, as well as with or without PASC diagnosis^74^. Patients classified as “stable” (Cluster2) were not taken into account, since representing an intermediate group with potential heterogeneity in its longer-term evolution, whereas clusters 1 and 3 were already identified with significant variations in their clinical parameters at the time of the last sampling. Then, all possible ratios were used as a threshold to define two groups: a high ratio group and a low ratio group. For each threshold, a Fisher’s exact test was conducted to determine the odds ratio (OR) of belonging to either cluster 3 or cluster 1, based on whether the participant fell into the high-ratio or low-ratio group. Fisher’s exact test was chosen due to the small sample size and the categorical nature of the data per sorted PBMC phenotype, ensuring accurate analysis of group differences.

The threshold giving the highest OR with p-value lower than 0.05 was selected as a good candidate to discriminate cluster 3 and cluster 1 patients in all analyses. The positive likelihood ratio (LR+) was also computed using this threshold. The positive likelihood ratio represents the likelihood of having an HERV-W *ENV* reads ratio in aMBc versus all other phenotypes above the threshold, therefore corresponding the high-ratio group for patients in cluster 3 compared to the likelihood of being in the high-ratio group for patients in cluster 1. The analysis was done using a custom R script.

### 2.3 Cells for transfection experiments

Human embryonic kidney (HEK) 293T cells (ATCC, CRL-3216) were grown in DMEM (Gibco, 41965-039) supplemented with 10 % of heat inactivated (30 min at 56°C) fetal bovine serum (FBS) (ATCC, 30-2020) and 2 mM L-Glutamine (ThermoFisher Scientific, 25030024) in 5% CO_2_ incubator at 37°C and were tested negative for mycoplasma spp.

### 2.4 Transfection

293T cells were cultured in 6-well plates (2,5.10^5^ cells/well) or in 8-well Nunc® Lab-Tek Chamber slide system (Merck, C7182) (5.10^3^ cells/well) precoated with Matrigel® (Corning, 354277). 24 hours after seeding, cells were transfected with 3 µg (6-well plates) or 0.3 µg (Lab-Tek chamber) of plasmid using Lipofectamine 2000 transfection reagent (Invitrogen, 11668-019). Three plasmids were transfected: pCMV-HERV-W ENV encompassing the complete ORF of pHERV-W ENV (HERV-W ENV clone, 542 amino acids GenBank no. AF331500.1) followed by U3R sequence, pCMV-HERV-W ENV-39STOP containing a single nucleotidic substitution introducing a STOP codon instead of the Tryptophan at amino acid position 39 (GeNeuro) or pCMV-Empty vector which shares the same backbone as the two previous plasmids but does not encode any protein. Plasmids and inserts are described in **supplementary Fig. 1.**

### 2.5 Treatment with nonsense mutations readthrough activity molecules

2 hours after transfection, 2,6-Diaminopurine (Merck, 247847-1G) previously resuspended at 50 mM in DMSO, G418 disulfate salt previously resuspended at 144 mM in sterile water (Merck, A1720) or Clitocine (CliniSciences, HY-118341, in DMSO) were added to the culture medium.

DMSO treatment condition is determined by the volume used for the highest dose of 2,6-Diaminopurine or Clitocine used in the experiment presented.

### 2.6 Immunofluorescence microscopy (IF)

48 hours post transfection, cells were washed once with 200 µL of Phosphate-Buffered Saline 1X (prepared from PBS 10X pH 7.4, Gibco, 70011-044) and fixed in 4% paraformaldehyde solution (Alfa Aesar, J61899) for 5 minutes. Cells were further washed three times with 200 µL of PBS 1X and permeabilized with 200 µL of PBS 1X supplemented with 0.2 % Tween 20 (Merck, P7949) for 15 minutes. After 30 minutes of incubation in 200 µL of blocking solution made with PBS 1X, 0.2% Tween 20 and 2.5 % horse serum (ATCC, 30-2040), cells were incubated with 50 µg/mL of anti-HERV-W ENV (GeNeuro, GN_mAb_Env02, murine antibody) diluted in blocking solution during 1 hour. After 3 washes with 200 µL of PBS 1X, cells were incubated for 1 hour with 1 µg/mL Alexa Fluor 488 goat anti-mouse IgG antibody (Invitrogen, A11029) and 0.1 µg/mL DAPI (Invitrogen, D3571) diluted in blocking solution. After 3 washes with 200 µL of PBS 1X, cells were mounted using Fluoromount-G mounting medium (Southern Biotech, 0100-01). Pictures were acquired on NIKON Eclipse TS2R microscope and analyzed on ImageJ software.

### 2.7 Protein extraction

48 hours post transfection, cells were harvested and lyzed in RIPA 1X buffer (prepared from RIPA 10X, Millipore, 20-188) supplemented with 1% Fos-Choline-16 (Anatrace, F316S 1 GM) and protease inhibitor cocktail (Roche, 04693132001). Lysates were incubated for 2 hours at 25°C with gentle agitation (120 rpm). After 10 minutes of centrifugation at 10,000 g and 4°C, supernatants were collected. Total protein amount was evaluated using Pierce™ 660 nm Protein Assay Reagent (Thermo Scientific, 22660). For some experiments, to improve HERV-W ENV hexamer extraction, supernatants previously collected were diluted 2 times in Solubilization buffer (part of Mem-PER Plus Membrane Protein Extraction Kit, Thermo Scientific, 89842), and incubated for 10 minutes at room temperature.

### 2.8 Automated Capillary Western Blot detection of HERV-W ENV

HERV-W ENV antigen detection was analyzed on the Wes device using Simple Western technology an automated capillary-based size sorting and immunolabeling system (ProteinSimple™). All procedures were performed with manufacturer’s reagents according to their manual. Briefly, 1500 µg/mL of protein lysate was mixed with fluorescent master mix and heated at 95°C for 5 minutes. The samples, blocking reagent, wash buffer, 20 µg/mL of primary antibody anti-HERV-W ENV (GeNeuro, GN_mAb_Env02, murine antibody), secondary anti-mouse HRP-coupled antibody (ProteinSimple, 042-205) and chemiluminescent substrate were dispensed into microplate. Protein samples were loaded into individual capillaries on a 25 capillary cartridge (12-230 kDa or 66-440 kDa separation matrix) provided by the manufacturer. Protein separation and immunodetection was performed automatically on individual capillaries using default settings. The global signal (AUC) containing the glycosylated monomer HERV-W ENV (electrophoregram peak about 120 kDa) and the hexamer HERV-W ENV (electrophoregram peak about 440 kDa) were both measured using Compass software for Wes device (ProteinSimple/Biotechne).

### 2.9 Quantitative RT-PCR (RT-qPCR)

48 hours post transfection, cells were harvested, and total RNA was extracted using NucleoSpin RNA Mini Kit (Macherey Nagel, 740955) according to the manufacturer’s protocol.

200 ng of DNase-treated RNA were reverse-transcribed into cDNA using iScript cDNA Synthesis Kit (Bio-Rad, 1708891) according to the manufacturer’s protocol. An amount of 5 ng of initial RNA in RT reaction has been used to quantitatively evaluate the transcriptional level of HERV-W *ENV* gene by RT-qPCR. The assays were performed in a StepOnePlus instrument (Applied Biosystems) using Platinum SYBR Green (Invitrogen, 11744-500). The housekeeping Glyceraldehyde-3-phosphate dehydrogenase (GAPDH) was used to normalize the results.

The RT-qPCR was performed using following primers: HERV-W ENV forward primer “fwd” [5’-GTATGTCTGATGGGGGTGGAG-3’] and reverse primer “rev” [5‘-CTAGTCCTTTGTAGGGGCTAGAG-3’] ; GAPDH fwd [5’-CACCCACTCCTCCACCTTTGAC-3’] and rev [5’-GTCCACCACCCTGTTGCTGTAG-3’].

Each experiment was completed with a melting curve analysis to confirm the specificity of amplification and the lack of any non-specific product and primer dimer. The conditions of amplification are presented in the graphical representation below, as generated by the software of StepOnePlus platform.

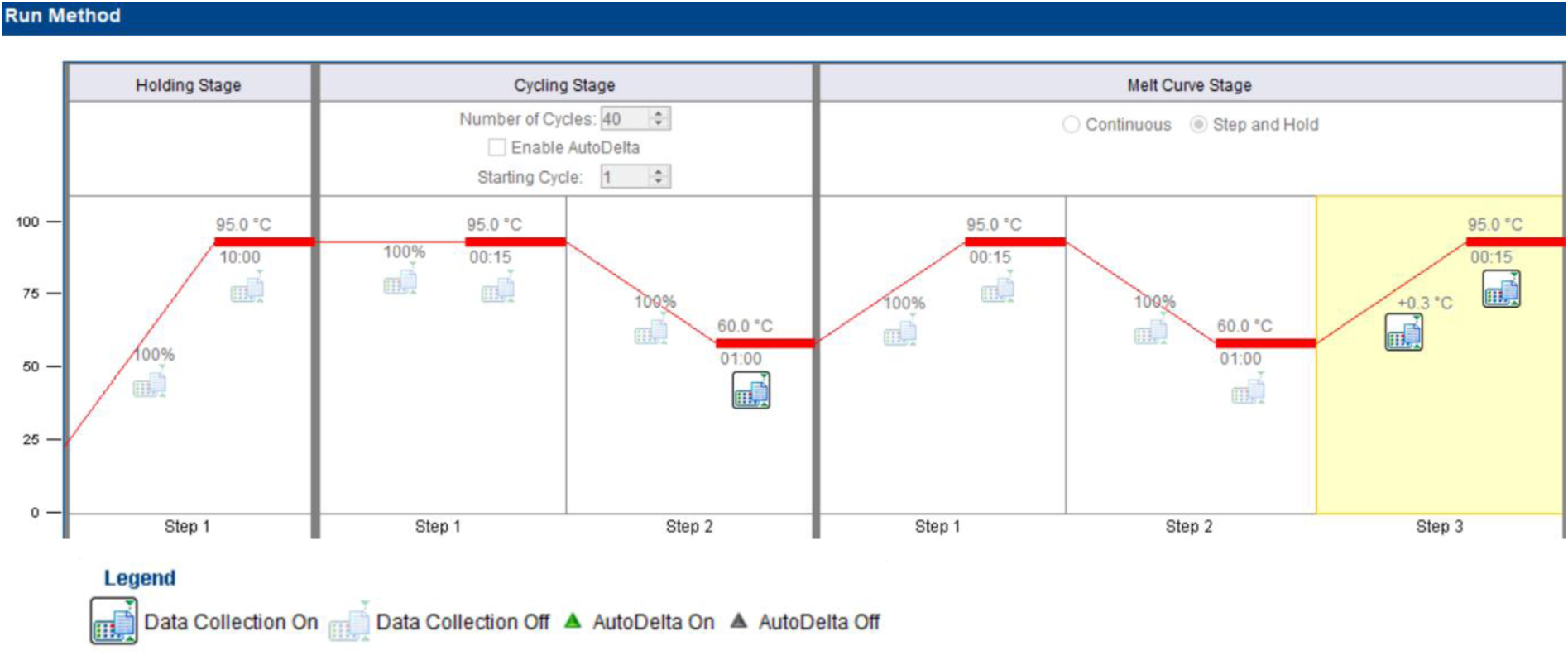

Quantification was performed using the threshold cycle (Ct) comparative method: the relative expression was calculated as follows: 2^-ΔCt (sample) -ΔCt (calibrator)]^ = 2^-ΔΔCt^, where ΔCt (sample) = [Ct (HERV-W *ENV*) – Ct (GAPDH)] and the ΔCt (calibrator) is the mean of ΔCt of “Empty vector” transfected cells for each treatment condition.

## 3. Results

Following previous studies on the PRESCO cohort^74^, a first analysis was performed on the samples from which RNA seq data were available over the successive visits for clinical follow-up, from visit 1 at early COVID-19 diagnosis to visit 5 at 3 months post-infection. Reads from all PBMC phenotypes aligning to HERV-W genomic sequences (HERV-W reference sequences in Supplementary information) were extracted and categorized per gene according to the protocol illustrated in **supplementary Fig. 2**.

The number of reads were first analyzed per million of total RNA seq reads pooled from all PBMC phenotypes. As illustrated in **Fig. 2A** and detailed in **supplementary Fig. 3**, the number of reads mapping to different genes or regions of HERV-W sequence varied with the PBMC phenotype and the post-infectious diagnosis of Post-acute COVID sequelae (PASC/long COVID). It clearly appeared that, when comparing samples from patients with PASC versus those with no PASC, the most elevated number of reads mapped to the ENV-U3R region in atypical memory B cells (aMBc). Reads showed a good coverage of the HERV-W *ENV* gene followed by the U3R sequence with highly homologous reads (end-to-end alignment), which corresponds to the expected mRNA since the poly-adenylation signal resides in the 3’ U3 sequences. No such level and significant difference was seen for the other HERV-W genes (gag, pol) or 5’LTR/RU5 sequences, though occasional and isolated differences between PASC and no-PASC samples were found in certain PBMC phenotypes for HERV-W pol, gag and RU5 sequences. These were associated with significantly elevated *ENV*-U3R reads in unswitched naïve B cells (BnUS) and central memory CD4 T cells (T4Cm), but with lower significance and relatively lower reads frequency (Cf. **supplementary Fig. 3**). Thus, HERV-W sequences with dominant ENV-encoding RNAseq reads appeared to be differentially expressed in three PBMC phenotypes versus others when comparing PASC and no-PASC patients from visit 1 and 2 during the early SARS-CoV-2 infection until visit 5, after COVID-19 infectious phase.

**Figure 2:**
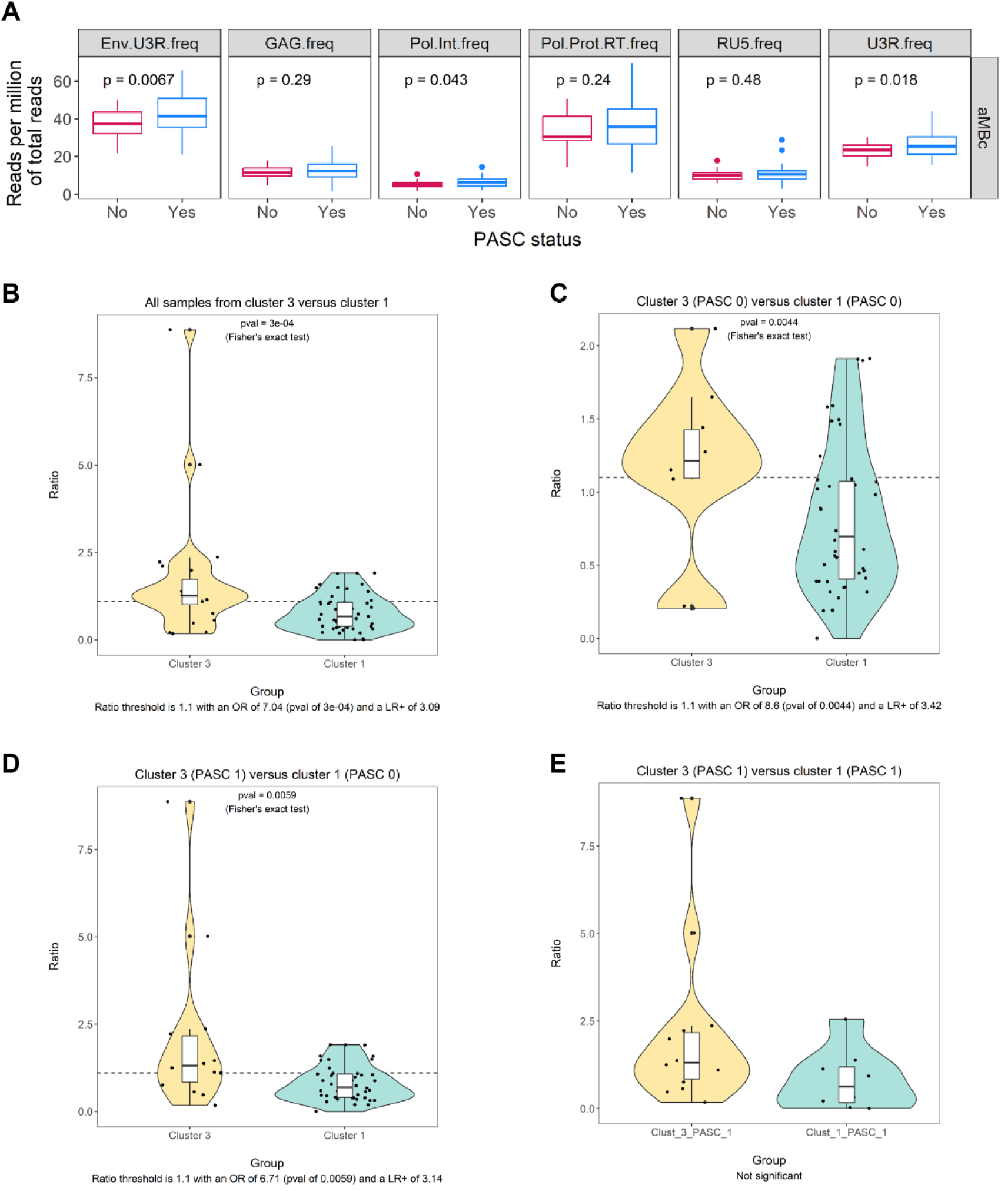
**A: Number of reads mapping to different genes or regions of HERV-W sequence in atypical memory B-cell PBMC phenotype (aMBc) from patients with or without Post-acute COVID sequelae.** The RNAseq reads of aMBc from earliest visit 1 at COVID-19 diagnosis to the last visit 5 post-infection at PASC diagnosis (CDC criteria) were pooled per patient and compared between those diagnosed with PASC (Long COVID) versus no PASC.Reads were mapped to HERV-W sequences (Cf. Fig. 1). Env-U3R (expected mRNA encoding the envelope protein), gag gene (GAG), pol gene region encoding the integrase (pol-int.) or the protease and reverse-transcriptase region (pol-Prot.RT), 5’ LTR (RU5) and 3’ LTR (U3R) sequences. **B-E:** Ratio of reads mapping Env-U3R between aMBc cells and all other phenotypes, in samples from patients with early SARS-CoV-2 infection: **2B.** Comparison of *ENV*-U3R reads ratio between all samples from cluster 3 versus all those from cluster 1. **2C.** Comparison of samples from patients without PASC, from cluster 3 versus patients from cluster 1. **2D.** Comparison of samples from patients with PASC in cluster 3 versus those with no PASC in cluster 1 taken as controls without PASC. **2E.** Comparison of all samples from patients later diagnosed with PASC according to CDC criteria, from cluster 3 versus cluster 1.

Globally, the RNAseq reads from pooled PBMC phenotypes showed a good coverage of *ENV-U3R* sequences, though relatively more abundant reads aligned to *ENV* 3’ end. This was even increased for the U3R sequence, which may result from the repeated sequences in this region (from retroviral Long-terminal repeat-LTR) but may also indicate variable thermodynamics in reads amplification between regions of the *ENV*-U3R sequence.

This prompted us to further seek potentially significant differences from a higher number of samples with RNAseq reads analyzed at visit 1 and 2 (n=97), while only few samples were made available with data from visit 5 (n=6). Among them, the PRESCO cohort comprised 80 patients with available RNAseq data from blood samples drawn during early COVID-19 (Visits 1 and 2) along with clinical follow up allowing both PASC diagnosis and retrospective analysis of the number and severity of COVID-19 symptoms. Patients’ clinical evolution and symptoms until last visit had made it possible to identify three clusters of patients irrespective of PASC diagnosis^74^: one of patients who spontaneously improved (cluster 1), one of patients with persisting but stable symptoms (cluster 2) and one of patients with an increased number and worsening symptoms (cluster 3). As expected, most patients diagnosed with PASC were diagnosed in cluster 3, representing most cases with numerous symptoms.

The analysis of the relative frequency of reads in RNA from FACS-sorted PBMC phenotypes from all patients at visit 1 and 2 showed that the most frequent HERV-W reads were still mapped to *ENV*-U3R sequences. They were then significantly detected in PASC or patients from cluster 3, within aMBc phenotype, which could be differentiated from all other sorted phenotypes. BnUs B and T4cm showed no difference, unlike previous analyses in which they had rather low frequency of reads but significantly different in PASC versus no-PASC when considering reads from post-infection visit 5. The reads aligning with *ENV* sequence only or *ENV*-U3R sequences also revealed comparable in aMBc. We therefore focused on the ratio of *ENV*-U3R reads in aMBc versus the pooled reads from all other PBMC phenotypes. This represented a ratio between an early *ENV*-expressing PBMC phenotype with potential HERV-W transcription, versus all others. This ratio also provided intra-sample normalization for comparison between samples from all patients. In the following analyses, samples from cluster 2 were not included, since representing an intermediate cohort that may require longer term follow-up to differentiate clinically defined evolution toward improved or worsened symptoms and their number.

In **Fig. 2B**, the comparison of *ENV* -U3R reads ratio between all samples from cluster 3 versus those from cluster 1 showed a quite complete differentiation between the clusters with a threshold at 1.1, an OR of 7.04 and a LR+ of 3.09. The statistical difference of *ENV* -reads between the two clusters was highly significant (p= 3e−04; Fischer exact test).

**Fig. 2C** compared patients from clusters 3 and 1, all of which were not later diagnosed with PASC and showed a very good discrimination between clusters. A threshold at 1.1 with an OR of 8.6 (p=0.0044; Fischer exact test) and a LR+ of 3.42, were found. Given that these samples were obtained during initial COVID-19, this difference should be due to the diverging evolution of symptoms and disease severity of the SARS-CoV-2 infection period between cluster 1 and cluster 3. This is consistent with HERV-W envelope RNA and protein expression in peripheral blood lymphocytes already described to be predicting, and associated with, pejorative disease evolution in early and late infection phases of COVID-19 across pandemic waves with various SARS-CoV-2 variants^17,70^.

**Fig. 2D** compared patients with PASC in cluster 3 versus those with no PASC in cluster 1, taken as controls without diagnosed PASC at Visit 5 and without persisting symptoms from acute COVID-19. These were not taken into account since persisting symptoms in cluster 1 may not be fully discriminant compared to PASC criteria. A very significant difference was confirmed to link PASC to the early detection of an elevated ratio of reads mapping to HERV-W *ENV*-U3R: Ratio threshold at 1.1 with an OR of 6.71 (p=0.0059; Fischer exact test) and a LR+ of 3.14. This corresponds to the detection of RNA transcription over the threshold in aMBc from the early days of COVID-19 in patients later diagnosed with PASC, with elevated values in few cases.

**Fig. 2E** similarly compared patients later diagnosed with PASC according to CDC criteria from cluster 3 versus PASC patients from cluster 1 and revealed no significant difference between them. This suggested a stronger association of *ENV*-U3R RNA expression with PASC than with the global clinical evolution considered to define the present clusters. A better discrimination provided by RNAseq data could thus be relevant when few PASC patients are found in the clinically defined cluster 1, though most of them were in cluster 3.

Further to these results showing HERV-W *ENV* transcriptional activity with reads mapping specifically to HERV-W sequences cloned from retrovirus-like particles of MS cell cultures and body fluids (originally named MSRV-*ENV*)^65,68^, the best possible alignment on human chromosomes still corresponded to with the previously described ERVWE2 copy^78^ that had been shown to encode a truncated protein^77^.

As shown in **Fig. 3A**, the reference HERV-W *ENV* sequence cloned from MS (GenBank ID: AF331500.1) is best aligned to a region in chromosome Xq22.3 (chrX: 106295765-106297364, with 1600/1629 aligned nucleotides and 98,5 % identity. This region corresponds to Homo sapiens RNA binding motif protein 41 (RBM41), Genbank sequence ID NG_016385.2. HERV-W *ENV* sequence is shown to correspond to the reverse strand of the RNA binding motif protein (RBM41) gene, starting from the end of intron 6 and ending within the beginning of exon 7. The RBM41 locus and the neighboring genes are presented in **Fig. 3B**, whereas **Fig. 3C** presents the positions of the 6 introns and 7 exons of RBM41 gene, all of which are used to generate alternatively spliced RNAs. The aligned nucleotide sequences also show that the entire ENV-U3R mRNA sequence shows 1936/2012 (96.22%) identities from Chromosome X positions 107052123-107054134. Thus, in **Fig. 4**, the complete HERV-W ENV-U3R sequence is found in this locus and therefore can generate a transcript corresponding to the mRNA structure on which RNA seq reads of aMBc from COVID-19 patients who developed PASC or had pejorative evolution (cluster 3) were significantly mapped. Moreover, as highlighted in yellow, TATA boxes are present upstream of the HERV-W *ENV* orf within the RBM41 intron 6, but with an antisense orientation since presented in the 5’ region of the ATG trinucleotide from which the translation of the HERV-W envelope can start. TATA and CAAT regulatory elements, with CAP site and Poly A signal in HERV-W (previously named MSRV) U3R sequences, along with strong promotor activity of MS-derived U3R clones, have also been described^68^. The additional presence of TATA boxes in RBM41 intron 6, upstream from this HERV-W *ENV* locus is indicating that a transcription is made possible from this HERV gene copy, independently from RBM41 normal RNA expression. This can be initiated within the RBM41 intron 6 where the ATG and the major part of the 5’ region of HERV-W *ENV* sequence is found, followed by the 3’ region within the RBM41 exon 7 where the downstream U3R region comprising promoter and regulatory sequences is located.

**Figure 3:**
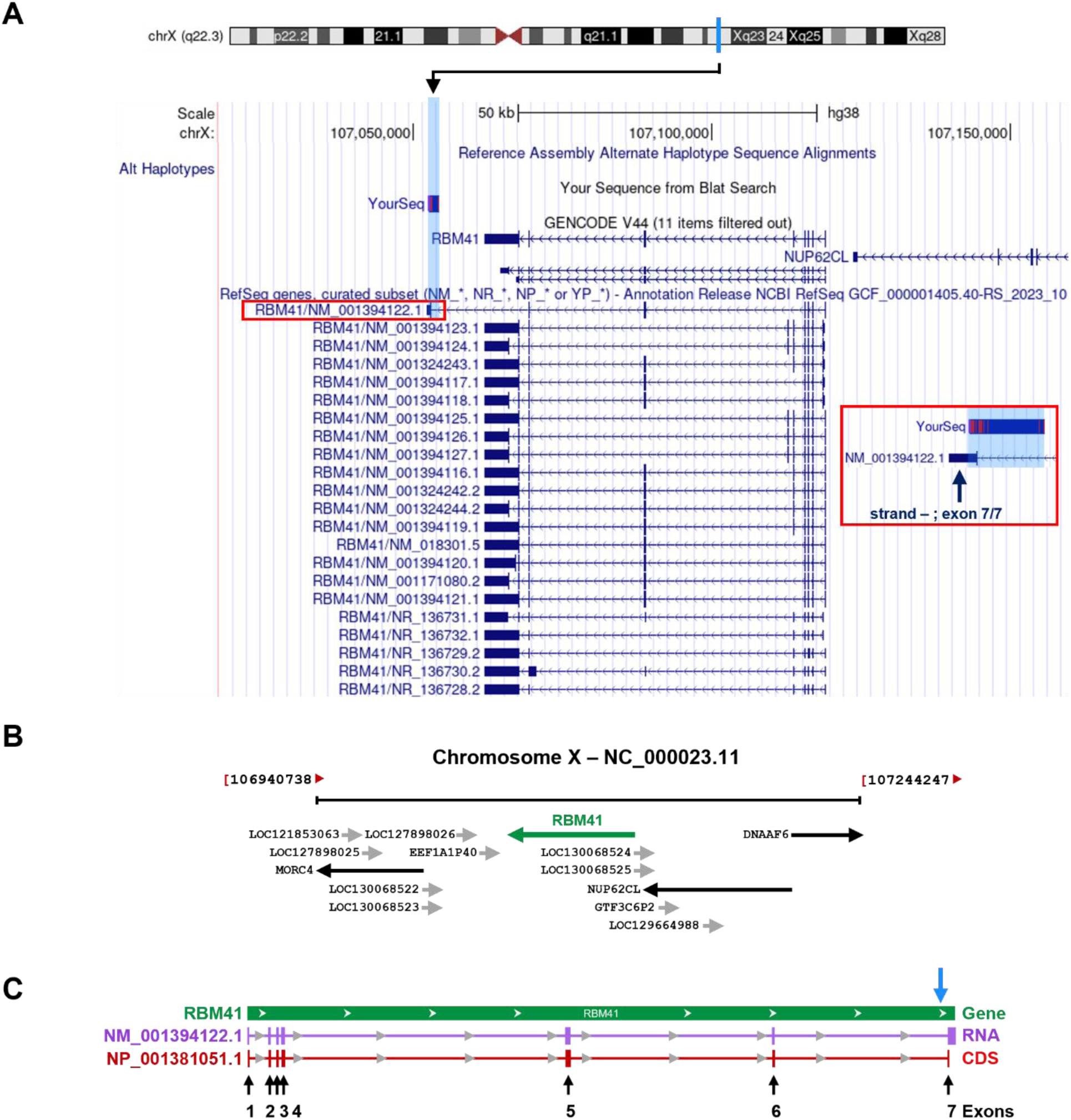
BLAT search of HERV-W ENV sequence using UCSC Genome Browser on Human (GRCh38/hg38). **A:** HERV-W ENV sequence is located (1600 nt aligned /1629 nt total ; 98,5 % identity) on the reverse strand of genomic chromosome X (chrX:107052926-107052997) on the region of RBM41 gene. The RNA reference RBM41/NM_001394122.1 boxed in red corresponds to the transcript variant 12 comprising exon 7 that contains the 3’ end of ENV-U3R sequence that is associated with the mRNA structure found to be expressed in COVID-19 aMBc. The protein isoform 12 expressed by this spliced RNA is predicted to be part of U12-type spliceosome complex. (provided by Alliance of Genome Resources, Apr 2022). The red square on the right shows, with blue background, the genomic region encompassing the HERV-W *ENV* sequence in an antisense orf starting in intron 6 and ending in exon 7. **B:** Localization of RBM41 gene on chromosome X, adapted from NCBI: https://www.ncbi.nlm.nih.gov/gene/55285. **C:** Genomic region (blue arrow indicates HERV-W ENV sequence alignment location), transcript and product of RBM41. RNA: mRNA-RNA binding motif protein 41, transcript variant 12. Protein: RNA-binding protein 41 isoform 12. Location: complement (107,052,102…107,118,822). CDS length: 1,095 nt. Protein length: 364 aa

**Fig. 4:**
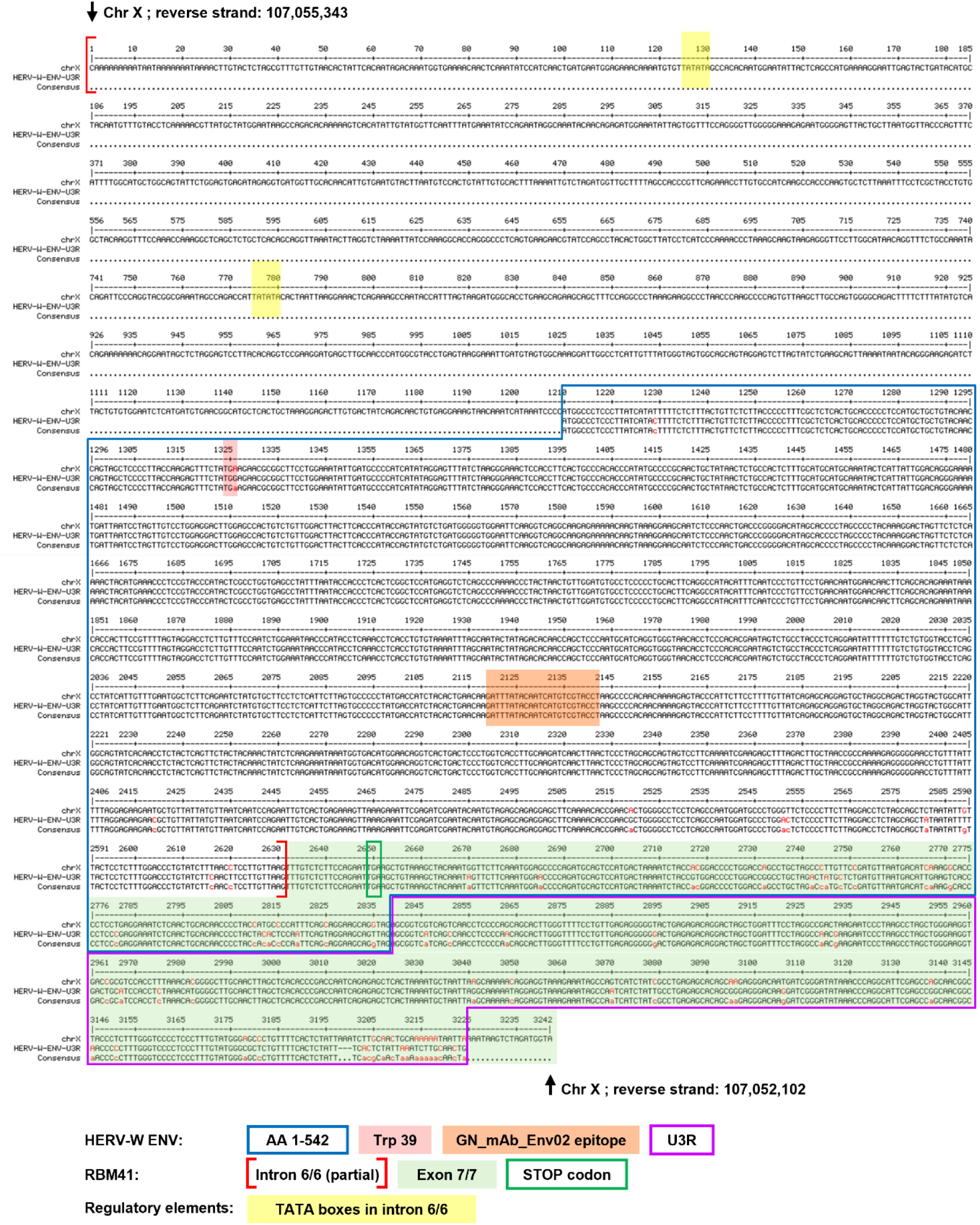
Aligned nucleotide sequences of Chr X portion 107052102 – 107055343 (reverse strand) with HERV-W *ENV*-U3R RNA sequence encoding the pathogenic HERV-W-ENV protein. TATA box motifs within chromosome X intron are highlighted in yellow; regulatory sequences in U3R, i.e., TATA boxes, CAAT element, Cap site and Poly A signal have already been described^68^. HERV-W ENV encoding nucleotides are boxed in light blue; nucleotides encoding tryptophane in position 39 of the HERV-W envelope protein and corresponding to the stop codon in chromosome X, are highlighted in light red; nucleotides encoding the epitope specifically recognized by Env02 monoclonal antibody used for immunofluorescence microscopy examination and immunocapillary western blot are highlighted in light orange; U3R sequences are boxed in purple. RBM41 intron 6 partial sequence are presented between red brackets in the aligned clone; RBM41 exon 7 sequence is highlighted in light green in the aligned clone ; the stop codon in the chromosome X corresponding to the terminal stop codon of RBM41orf for the isoform 12 is boxed in green.

The aligned proteins respectively encoded by the reference sequence on chromosome X, also named ERVWE2, and the HERV-W sequence, previously named MSRV, cloned and sequenced from an MS sample^41^ are shown in **supplementary Fig. 4** . A stop codon is interrupting the orf from the ERVWE2 locus and the corresponding RNA might be translated into a truncated protein from an ATG trinucleotide encoding a methionine downstream the position of tryptophane 39 as shown *in vitro*^77^. This in-frame stop codon within the ERVWE2 locus precluded the expression of a full-length protein with the specific biochemical and biophysical properties characterized in native HERV-W ENV antigen extracted from, e.g., MS brain lesions^20^. However, we recently understood that such stop codons are used for the regulation of protein expression at the ribosome level in procaryotes^87,88^ and that some mechanisms can promote ribosome readthrough in vertebrates^89–92^. Moreover, molecules were proposed for therapeutic strategies promoting such ribosome readthrough in human genetic diseases with stop codons impairing the production of physiological proteins^93–95^. Therefore, such ribosome readthrough might be possible for HERV genes in human cells. We then conceived experiments to find conditions that could provide proof of concept for a full length HERV-W ENV protein expressed from this ERVWE2 sequence in human cells, despite an in-frame stop codon.

To experimentally test this hypothesis, we generated a plasmid with the corresponding ERVWE2 stop codon in the HERV-W *ENV* orf. In parallel, we have used the HERV-W *ENV* plasmid encoding the complete envelope protein cloned from an MS sample and used to produce a recombinant protein (GenBank ID: AF331500.1)^41^. The constructs are schematically presented in **supplementary Fig. 1**. The stop codon in the HERV-W ENV was confirmed by plasmid sequencing. Of note, though the cleavage of the signal peptide is observed with full-length HERV-W envelope form plasmid expression, the Furin cleavage site is inactivated by mutations found in the original clones from MS^65,68^ and confirmed within the HERV-W ENV proteins extracted and analyzed from MS brain lesions^20^. The produced full-length envelope protein therefore no longer contains the propeptide, but still retains both surface (SU) and transmembrane ™ units.

HEK cells were transfected with either plasmid construct and with different molecules shown to have a possible effect on ribosome readthrough of in-frame codons, lysed and analyzed with Simple Western immunocapillary platform (Proteinsimple, CA, USA), as detailed in Materials and Methods. In **Fig. 5**, results from transfection in the presence of three molecules already shown to display such an effect in humans, 2,6-Diaminopurine (DAP)^95^, G418^96^ and clitocine^97^ are presented in parallel with results of the transfection in normal medium (no treatment). DMSO at the same concentration as used to solubilize 2,6-DAP or clitocine, was also compared as a SHAM control. As can be seen with the electrophoregram from the Simple Western immunocapillary analysis, a peak of protein immunolabelled with a specific anti-HERV-W ENV mouse monoclonal antibody (GN_mAb_Env02) migrating at the expected molecular weight of the mature (without propeptide) glycosylated HERV-W ENV monomer was seen in all conditions with the W ENV plasmid. The transfection with W ENV-39 STOP plasmid containing the stop codon showed no expression in normal medium, even under the form of a truncated protein with lower molecular weight as previously made possible *in vitro*^77^, which would have been possible from the position of the recognized epitope (Cf. **Fig. 4** and **supplementary Fig. 4**). No expression was either observed with W ENV-39 STOP plasmid in the presence of G418 or clitocine. However, in the presence of DAP, the expected peak of the mature and glycosylated complete HERV-W ENV monomer was observed with 100 µM DAP in the culture medium (**5D**). A dose-response experiment with DAP at increasing concentrations was then performed and, as seen in **5F**, an effect ranging from 50 µM to 150 µM with an optimal concentration of 100 µM DAP for the expression of HERV-W ENV protein was confirmed for the plasmid with the stop codon, along with good reproducibility between experiment duplicates. To confirm the analysis of the denatured protein from transfected cells as previously performed, immunocytology microscopy analysis was performed with the same monoclonal antibody (GN_mAb_Env02) using 50 µM concentration of DAP. In **5G**, immunofluorescent staining was observed in all conditions with the W ENV plasmid, whereas the same positivity and pattern of expression with strong labelling at the membrane level was only observed in the presence of DAP with the W ENV-39STOP plasmid.

**Fig. 5:**
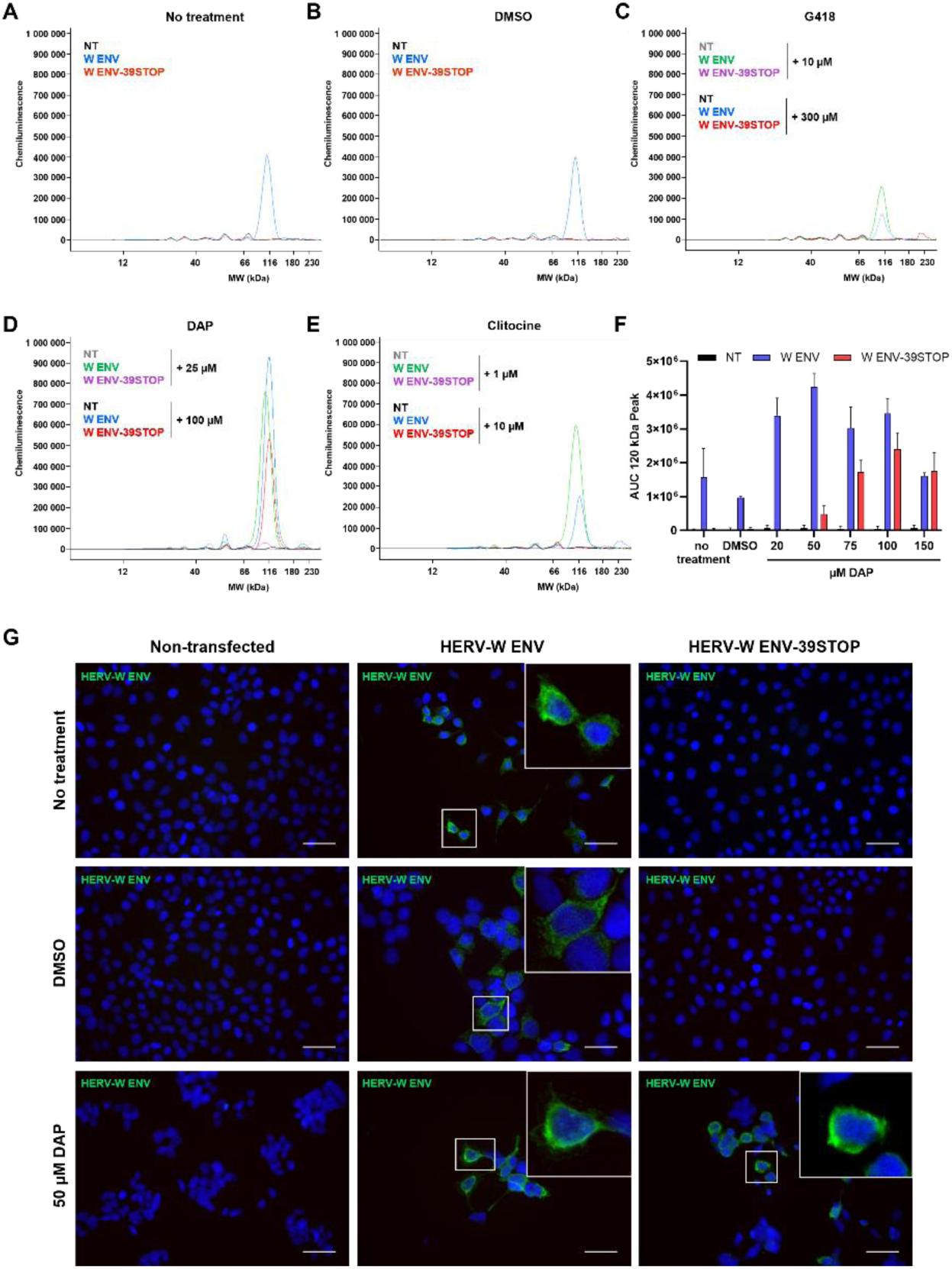
Identification of DAP as an efficient molecule to overcome stop codon in HERV-W ENV-39STOP construct. **A-F:** Non-transfected 293T cells, 293T cells transfected with the plasmid coding for HERV-W ENV or for HERV-W ENV-39STOP construct were treated with different molecules known for their nonsense mutation readthrough activity. 48 hours post transfection, total proteins were extracted and HERV-W ENV expression analyzed by Simple Western on 12-230 kDa size separation matrices. Non-treated cells (**A**), cells treated with DMSO (**B**), 10 or 300 µM G418 (**C**), 25 or 100 µM DAP (**D**), 1 or 10 µM clitocine (**E**) or DAP range from 20 to 150 µM (**F**). Results are presented as migration electrophoregrams (**A-E**) where the 120 kDa peak corresponds to full length HERV-W ENV glycosylated monomer expression or as quantitative interpretation based on calculation of the area under curve (AUC) of 120 kDa peak (F, mean+SD of 2 independent capillaries). **G:** Non-transfected 293T cells (left panel), 293T cells transfected with the plasmid coding for HERV-W ENV (middle panel) or for HERV-W ENV-39STOP construct (right panel) and untreated (top panel), treated with DMSO (middle panel) or with 50 µM DAP (bottom panel) were stained 48 hours post transfection using GN_mAb_Env02 antibody (green staining). DAPI was used to stain nuclei (blue staining). Scale bars = 15 µm.

Because the HERV-W ENV antigen extracted from MS brain lesions had revealed to be present *in vivo* with an oligomer conformation (hexamer) characterizing a secreted soluble form of this pathogenic expression and could be reproduced with the recombinant protein expressed from the present W ENV clone^20^, we looked for this high-molecular weight form on appropriate capillaries. Of note, this hexameric form resisted strong denaturing conditions used for the simple western capillary analysis and other western-blotting formats. In **Fig. 6A**, both HERV-W ENV monomer around 120 KDa and hexamer around 440 KDa could be detected in cells transfected with W ENV-39STOP plasmid and 75 µM DAP, similarly to what was obtained with the W ENV plasmid in normal medium. A dose-response analysis showed that the monomer was similarly detected with W ENV at 20 to 75 µM of DAP, but slightly increased compared to SHAM condition with DMSO. However, the decrease at 100 µM could indicate that some cell toxicity was encountered at this concentration. In these conditions, the monomer was increasingly expressed from 20 to 75 µM DAP with W ENV-39STOP plasmid and, though non-significant difference was seen at 100 µM, the mean + SD of 2 to 8 independent capillaries from 2 to 4 independent experiments was reduced (**6B**). The same dose-response analysis at the level of the hexamer molecular weight also revealed an increasing production from 20 to 75 µM DAP with W ENV-39STOP plasmid, whereas a potential cytotoxicity was confirmed with a clear decrease at 100 µM (**6C**). Finally, because differences were confirmed to result from factors acting at the translational level, we performed a study of the transcriptional levels to compare the RNA expressed from the different conditions of transfection with both W ENV and W ENV-39STOP plasmids. No significant difference between the transcriptional levels of each plasmid was observed in all conditions, thereby confirming that differences observed at the protein level are not influenced by the transcription of either sequence in transfected cells.

**Fig. 6:**
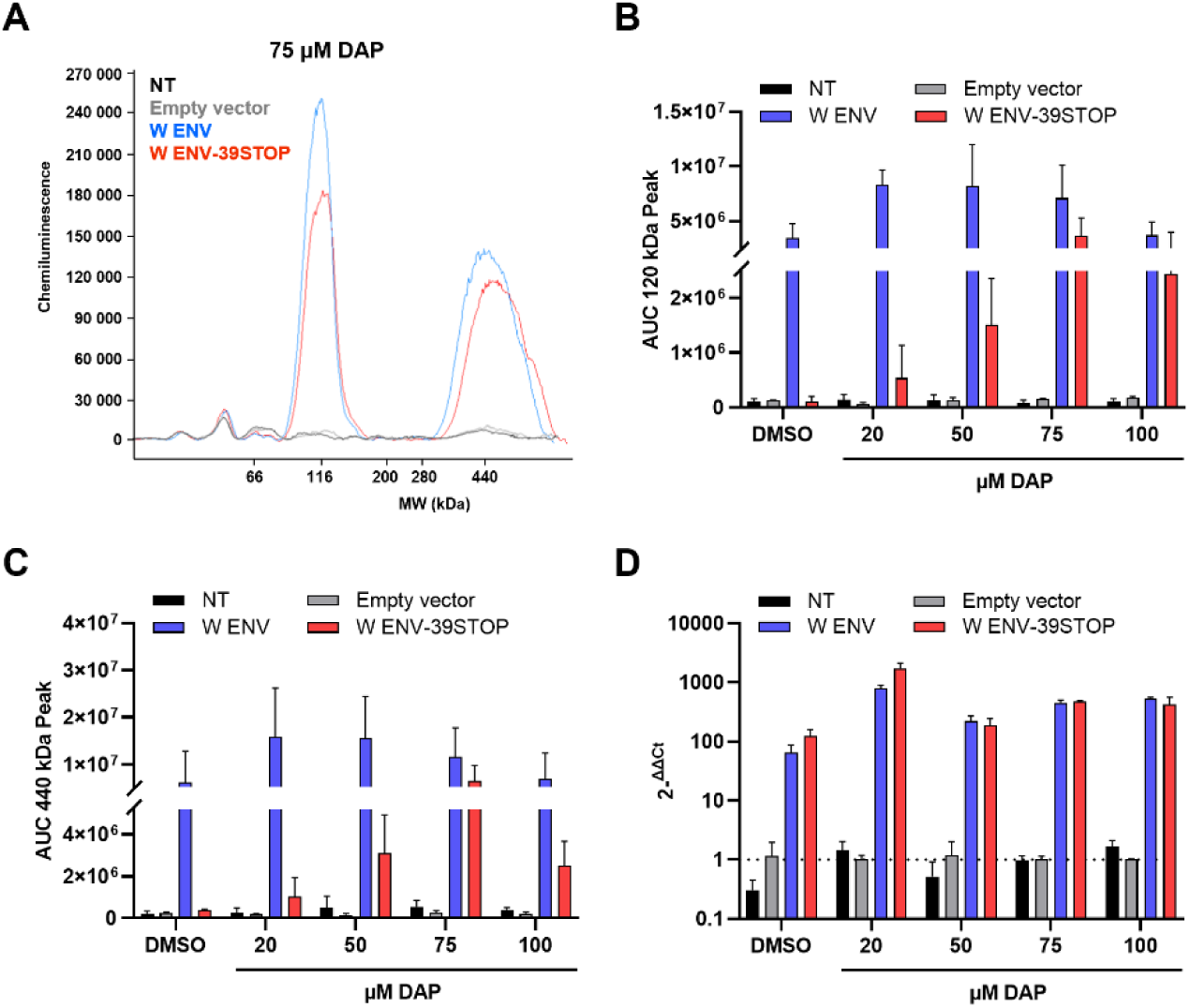
HERV-W ENV protein induced by stop codon readthrough can still form hexamer. Non-transfected 293T cells, 293T cells transfected with the empty vector or with the plasmid coding for HERV-W ENV or for HERV-W ENV-39STOP construct were treated with DMSO or DAP range from 20 to 100 µM. (**A-C**) 48 hours post transfection, total proteins were extracted, soluble fractions were incubated with a second buffer to improve hexamer extraction and HERV-W ENV expression analyzed by Simple Western on 66-440 kDa size separation matrices. Results are presented as migration electrophoregram (**A**, example of 75 µM DAP) where the 120 kDa peak corresponds to full length HERV-W ENV glycosylated monomer expression and the 440 kDa peak to the hexameric form of the protein or as quantitative interpretation based on calculation of the area under curve (AUC) of 120 kDa peak (**B**, mean+SD of 2 to 8 independent capillaries from 2 to 4 independent experiments) or 440 kDa peak (**C**, mean+SD of 2 to 8 independent capillaries from 2 to 4 independent experiments). (**D**) 48 hours post-transfection, cells were harvested and total RNA extracted. RT-qPCR was performed to determine the transcriptional level of HERV-W *ENV* gene for each transfection and DAP treatment condition. Results are presented as the mean+SD of 2^-ΔΔCt^ from three independent experiments.

## 4. Discussion

In a first part, this study completes the ones already performed on clinical, immunological and other multi-omics parameters from the PRESCO cohort^74–76^, here focusing on RNASeq data mapping the HERV-W sequences. The rationale for this study on patients relied upon the facts that HERV-W was previously shown to encode an immune- and neuro-pathogenic envelope protein^24,98^ whose expression in lymphoid cells and release as soluble antigen in peripheral blood (i) could be induced by SARS-CoV-2^17^ and (ii) correlated with COVID-19 severity and/or a subgroup of patients with long COVID (PASC)^72,73^.

The results from this RNAseq analysis, show that HERV-W RNA reads mapping to the envelope-encoding mRNA reference sequence, originally cloned from cultured cells or CSF of patients with multiple sclerosis (MS), were significantly elevated in atypical memory B cells (aMBc) of early SARS-CoV-2 infection in patients who were later diagnosed with Long COVID, but also in those who experienced worsened COVID-19 and late symptoms. Interestingly, B lymphocytes have long-been shown to be permissive for this pathogenic HERV-W expression in MS and other diseases^24,43,99–102^. It is therefore of the greatest interest to learn that B lymphocytes infected with Epstein-Barr virus (EBV) undergo differentiation into aMBc and that aMBc are pathogenically associated with numerous immune disorders^103^. Thus, HERV-W pathogenic expression induced in such B cells in “genetically susceptible” individuals, as seen with PBMC exposure to SARS-CoV-2^17^, is likely to be an early feature of related pathogenic post-infectious dynamics and appears to share common features between EBV and SARS-CoV-2 infections, though global post-infectious kinetics are expected to differ.

This early detection of specific RNA in aMBc is likely to provide a predictive biomarker of pejorative COVID-19, particularly in patients at risk or with undiagnosed comorbidities whatever the SARS-CoV-2 strains, vaccine or previous infection status. This is also consistent with peculiar findings on aMBc in COVID-19 showing the prominent expansion of aMBc in severe COVID-19^104^, suggested to represent one of the critical drivers in disease severity^105^, and associated with an impaired neutralization potency post third dose in the elderly vaccinated with an adenovirus-vectored vaccine but not observed with mRNA SARS-CoV-2 vaccines^106^. However, given the present public health burden caused by the post-COVID syndromes affecting millions of individuals worldwide and their continuous incidence despite the rising global immunity and the emergence of less pathogenic coronavirus strains, this predictive detection from early infection paves the way to therapeutic prevention with precision medicine targeting this pathogenic HERV-W expression. Such biomarker-based therapeutic prevention should be made possible using the Verily’s Immune Profiler platform ^76^ or adapted protocols to perform a similar quantification of the HERV-W ENV-encoding mRNA in aMBc from SARS-CoV-2 early infected patients. This is being studied with the W-ENV neutralizing IgG4 antibody, temelimab, currently evaluated in a phase II placebo-controlled clinical trial focusing on long-COVID/PASC patients with HERV-W ENV antigenemia (ClinicalTrials.gov ID NCT05497089). Consequently, the prevention of COVID-19 complications and, most of all, of post-COVID condition known as PASC or Long COVID remains a challenge that is likely to be addressed from early infection with a biomarker-based targeted strategy. Even if the persistent expression of W-ENV should not be relevant for all cases, it represents a major subgroup of patients with complex and highly heterogeneous post-COVID symptoms for which a biomarker-based precision medicine is made possible.

In a second part and, further to this early mRNA expression specifically observed in aMBc from COVID-19 patients, the present study re-addressed the question of the involved chromosomal HERV-W gene copy in the light of new knowledge on the regulation of mRNA at the ribosome level. SARS-CoV-2 was shown to induce HERV-W ENV protein expression in peripheral blood mononuclear cells (PBMC) from healthy blood donors maintained in normal cell-culture medium, with RT-qPCR detecting a peak of RNA transcription at 2h post-exposure and with detectable protein expression by immunocytology in macrophage and lymphocyte cells at 48h post-exposure. Apart from the fact that some (epi)genetic factors may explain why only about 20% of donor’s PBMC did express HERV-W ENV when exposed to SARS-CoV-2^17^, the present results confirming a complete mRNA sequence transcription in clinically worsening COVID-19 patients implicitly raised the question of the origin of this protein expression.

The results of the present study show for the first time that an HERV protein encoded by a gene with a stop codon can be expressed in human cells in presence of a specific “helper” molecule. Moreover, it confirmed that the reference HERV-W ENV sequence on which RNAseq reads from COVID-19 patients mapped, best aligned to the previously mentioned ERVWE2 locus on chromosome X^77–79^. This remains the only known chromosomal HERV-W copy of HERV-W envelope gene from the present databases with this nearly perfect nucleotide sequence identity. Therefore, the previously considered ERVWE2 impossibility to encode and be translated into the full-length protein with characteristics identified from MS brain lesions^20,34^, can now be understood to be untrue. This, since the nucleotide sequence with its in-frame stop codon has been shown to yield the full-length HERV-W ENV protein forming hexamers as shown for the MS-related protein *in vivo* and *in vitro*. So, though an early triggering event with delayed clinical emergence in MS is likely to be provided by an EBV infection that might have caused somatic rearrangements or mutations^80,102,107^ followed by “boosting” agents or by alternative triggering

infections^25,38^, the quite immediate induction of HERV-W ENV expression caused in lymphoid cells from healthy individuals by SARs-CoV-2 suggest a direct effect on an HERV-W copy as it is present in human chromosomes without delayed effects of potentially induced DNA rearrangements. Thus, the present results showing that a regulation of HERV mRNA expression with in-frame stop codon(s) can be achieved at the ribosome translation level in human cells as reported in other species and human conditions, elucidates the origin of the corresponding HERV-W ENV protein production, at least the one involved in COVID-19. Interestingly, all sequence motifs making it possible to induce the autonomous transcription of this HERV-W gene copy from chromosome X, independently from RBM41 gene transcription have now been identified. Thus, HERV-W ENV transcription can start from upstream TATA boxes in RBM41 intron 6 ending into its exon 7. The presence of HERV-W U3R regulatory sequences in ERVWE2 locus is allowing interactions with viral encoded or virus-induced molecules as previously reviewed^26^, but may also be involved in a potent (epi)genetic repression of its transcription in about 80% of “non-responding” individuals (versus about 20% of “responders”)^17^.

In conclusion, RNAseq data from COVID-19 patients along with the renewed analysis of ERVWE2 locus in chromosome X and the provided experimental proof of concept that its in-frame stop codon is not precluding the production of HERV-W ENV protein, elucidates a new possible mechanism at the ribosome translation level. This can occur when a “helper” molecule is present that makes a readthrough process possible for the corresponding mRNA and therefore suggests that SARS-COV-2 directly or indirectly provides this key factor. Moreover, as the activation of the ERVWE2 copy transcription is seen possible whenever epigenetically feasible, it also suggests that SARS-CoV-2 would first contribute to trigger regulatory elements leading to its autonomous transcription, while allowing its downstream translation. Studies on such mechanisms induced by SARS-COV-2 are now needed to further elucidate viral factors involved in such post-infectious HERV activation and expression.

## Data Availability

All data produced in the present study are available upon reasonable request to the authors

## Acknowledgements

This study was supported by Geneuro-Innovation SAS, France.

