## Supplementary material for "An HERV-W *ENV* transcription in atypical memory B cells linked to COVID-19 evolution and risk for long COVID can express the encoded protein from a ribosome readthrough of mRNA from chromosome X"

**Supplementary data :**

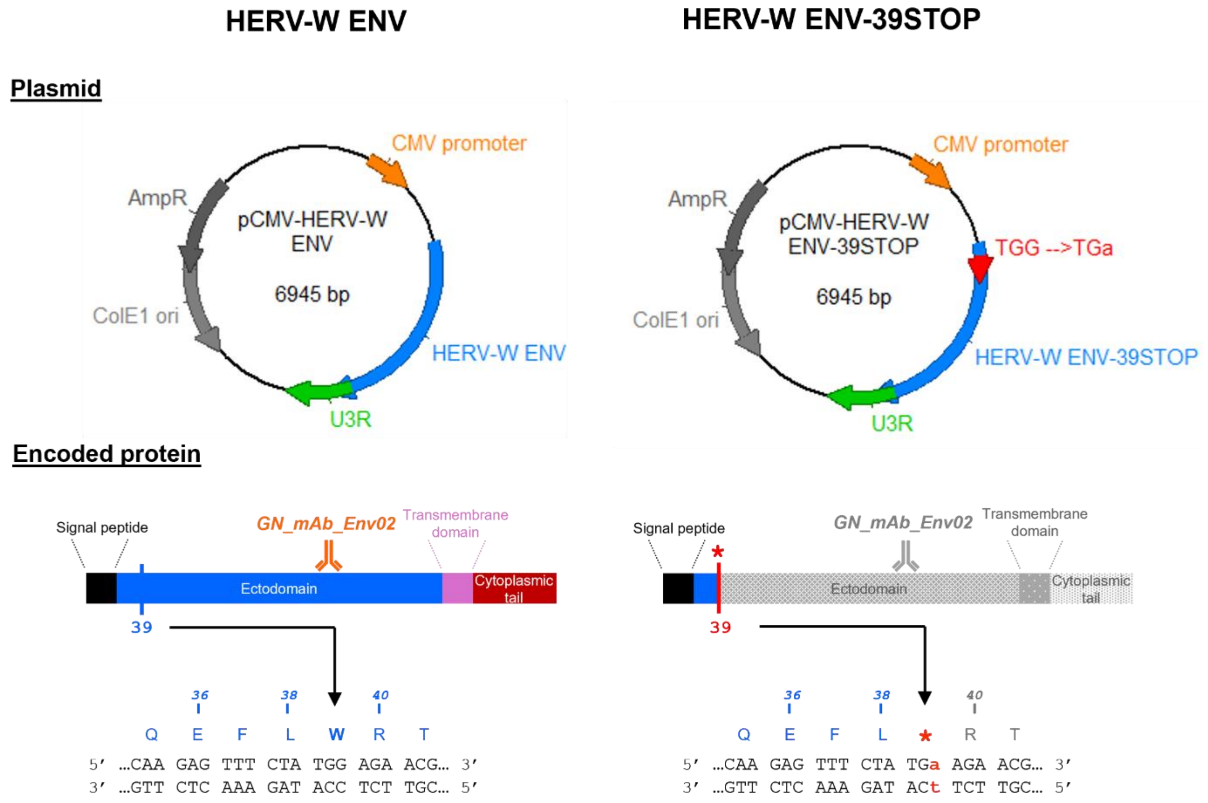


**Supplementary Figure 1: Schematic presentation of HERV-W ENV and HERV-W ENV-39STOP constructs for transfection experiments**

**Top:** pCMV-HERV-W ENV and pCMV-HERV-W ENV-39STOP plasmids**.** HERV-W ENV and HERV-W ENV-39STOP sequences were followed by the U3R sequences, as cloned from MS samples and as found to be highly similar in the chromosome X copy, antisense within RBM41 exon 7.

**Bottom:** HERV-W ENV and HERV-W ENV-39STOP proteins encoded by the plasmids. Epitope specifically recognized by Env02 monoclonal antibody used for immunofluorescence microscopy examination and immunocapillary western blot is symbolized by orange antibody on HERV-W ENV ectodomain and grey antibody on HERV-W ENV-39STOP construct.

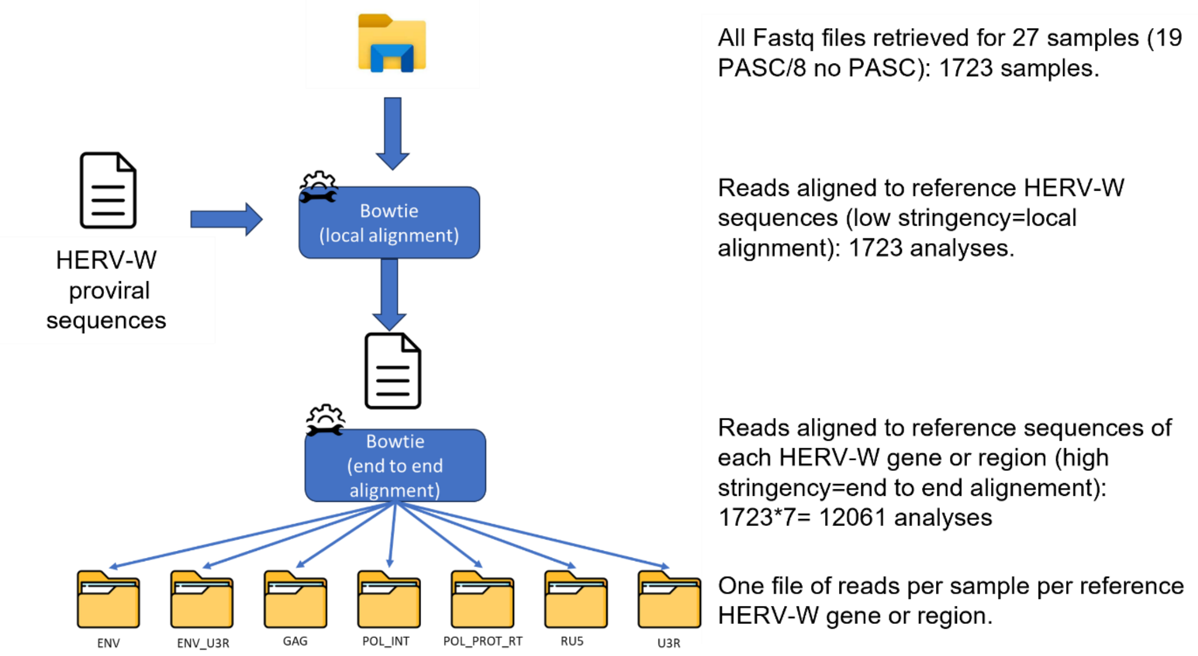


**Supplementary Figure 2: RNA seq reads from all cell-types mapped to *ENV*-U3R HERV-W genomic sequence**

Frequency of pooled RNAseq reads mapping end-to-end with the expected ENV-U3R RNA. RNAseq reads of representative patients with follow-up samples and RNAseq data pooled from visit 1 to 5. The frequency of mapped reads is expressed as the number of reads per million of pooled RNA seq reads of all PBMC phenotypes.

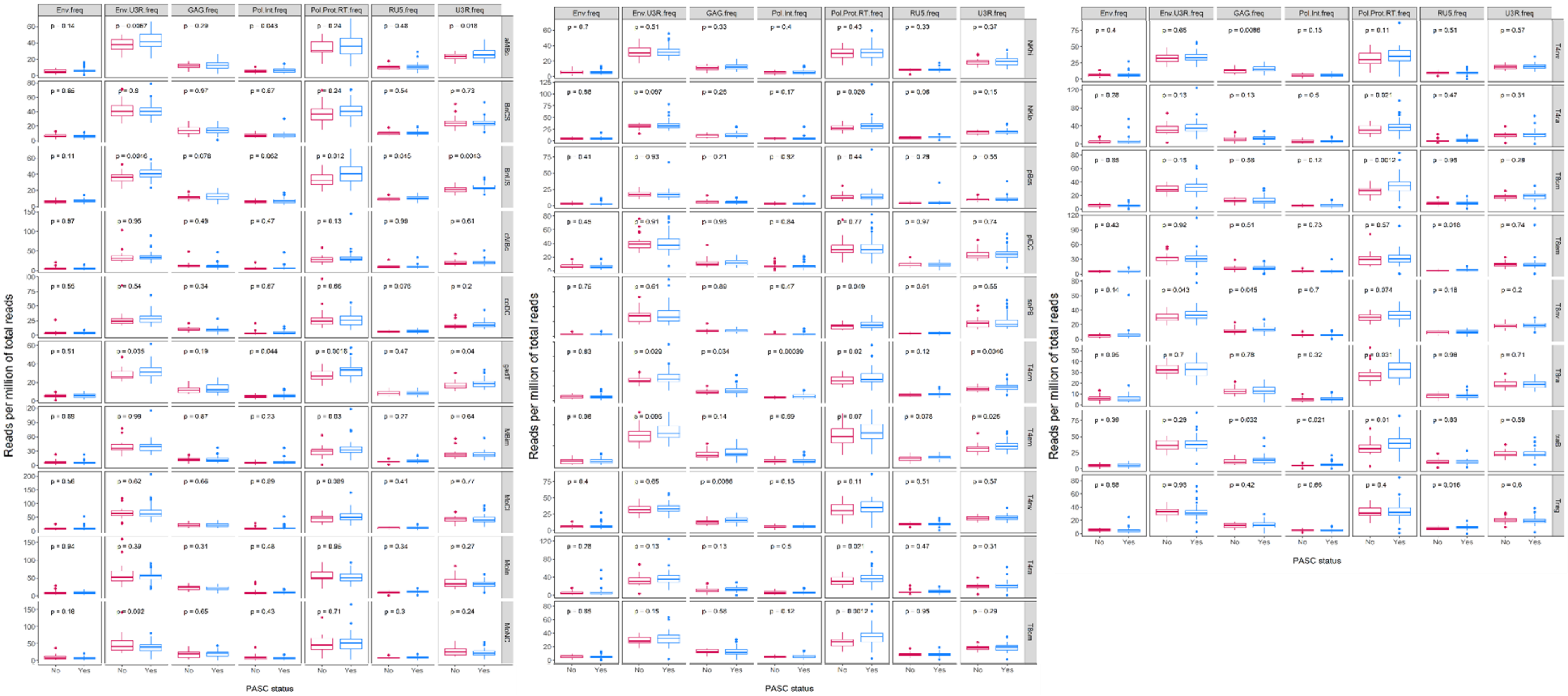


**Supplementary Figure 3: Detail of number of reads mapping to different genes or regions of HERV-W sequence in PBMC.**

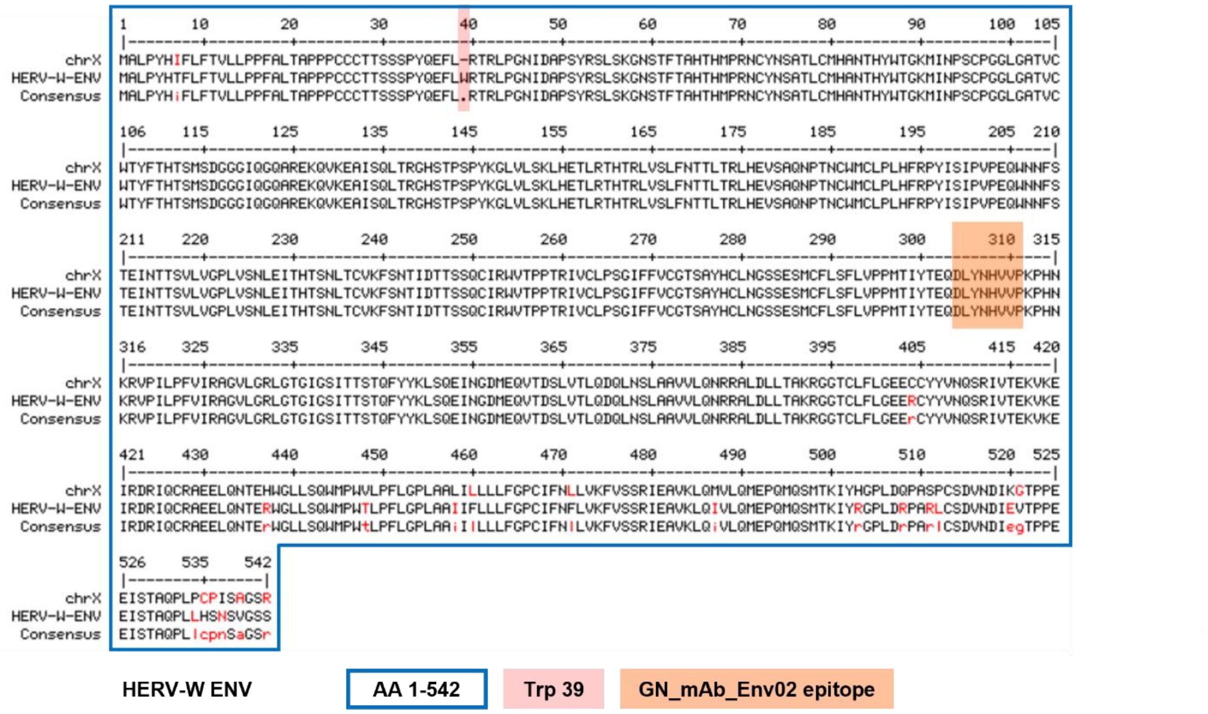

**Supplementary Figure 4: Amino acids alignment between HERV-W-ENV and the nearly identical protein encoded by the mapped sequence on Chr X.**

Proteins respectively encoded by the reference sequence on chromosome X, also named ERVWE2, and the HERV-W sequence, also name MSRV-*ENV* cloned from an MS sample and used to produce recombinant HERV-W envelope protein (GenBank ID: AF331500.1). As highlighted in light red, a stop codon is interrupting the orf from the ERVWE2 loci where a tryptophane is encoded at position 39 of the amino acid sequence. Amino acids of the identical epitope specifically recognized by Env02 monoclonal antibody used for immunofluorescence microscopy examination and immunocapillary western blot are highlighted in light orange.

**Supplementary data folder and document:**
-Reference HERV-W dataset file (refseq)

-Example of iterative code for data retrieval from the PRESCO RNAseq dataset :
Tools used for the analysis

bowtie2 v 2.2.3
fastqc v0.11.9
multiqc v1.14.dev0

Command line used for each sample analyzed

Concat all references in one Fasta file : All_references.fasta
ENV.fasta
ENV_U3R.fasta
GAG.fasta
HERW.fasta
POL_INT.fasta
POL_PROT_RT.fasta
RU5.fasta
U3.fasta
U3R.fasta
U5.fasta

```
# Build of bowtie reference index for each reference
# this command line was done for each fasta file of reference used during the analysis (11 times)
e.g bowtie2-build  ENV.fasta ENV
```

```
# Retrieve all potential reads of interest using local alignement mode
# This command was performed for all sample analyzed (1923 time for first analysis and 2210 times for second analysis)
bowtie2 -p 18 --local -k 1 -N 1 -L 20 -i S,1,0.75 --ignore-quals --ma 4 --mp 12,12 --score-min G,-20,34 -x All_references -U SampleX_R1.fastq.gz,SampleX_R2.fastq.gz --no-unal --al Sample_Extracted_reads.fastq
```

```
# Based on reads extract we identify by a more stringent analysis reads that could be associated to each target (exemple for ENV)
bowtie2 -p 6 --end-to-end -k 1 --very-sensitive --ignore-quals -x ENV -U Sample_Extracted_reads.fastq --no-unal --al Sample_ENV_reads.fastq
```

```
#fastqc analysis to determine easlily the number of reads per files (this is done in each folder that contains all the reads extracted)
fastqc -t 12 -o Quality/ *.fastq
```

```
# multiqc analysis to retrieve all fastqc information in one report (Quality folder in same as previous command)
```

multiqc -o Quality/MutiQCResults/ Quality/

All the previous commands were done for each sample and for each targets

We obtain 2 folders one for the first Analysis (1923 samples) and for the second analysis (2649 samples) that contain the following folder
ENV
ENV_U3R
GAG
POL_INT
POL_PROT_RT
RU5
U3R

Then we used the following java code to retrieve all read numbers information for each samples
As input we used results of multiQC analysis results for each target analysis, raw data analysis.
We used also a text file "Patient_info.txt" that contain for each patient of the first analysis the PACS information. 'Yes' or 'No'

```
import java.io.BufferedReader;
import java.io.BufferedWriter;
import java.io.FileReader;
import java.io.FileWriter;
import java.io.IOException;
import java.util.HashMap;
import java.util.List;
import java.util.Map;
import java.util.Set;

public class FastqcAnalysisToRetreiveReadsVerily
{


	public static void main(String[] args)
	{


// ANLAYSE 1
//		String fullReadsInfosPath ="Raw_Data_quality_Analysis/MutiQCResults/multiqc_data/multiqc_fastqc.txt";
//		String fullExtractReadsInfosPath ="Extract_Reads/Quality/MutiQCResults/multiqc_data/multiqc_fastqc.txt";
//		String ENVReadsInfosPath ="Specific_Reads_Analysis/ENV/Quality/MutiQCResults/multiqc_data/multiqc_fastqc.txt";
```

```
//		String ENV_U3RReadsInfosPath
="Specific_Reads_Analysis/ENV_U3R/Quality/MutiQCResults/multiqc_data/multiqc_fastqc.txt"
;
//		String GAGReadsInfosPath
="Specific_Reads_Analysis/GAG/Quality/MutiQCResults/multiqc_data/multiqc_fastqc.txt";
//		String POL_INTReadsInfosPath
="Specific_Reads_Analysis/POL_INT/Quality/MutiQCResults/multiqc_data/multiqc_fastqc.txt";
//		String POL_PROT_RTReadsInfosPath
="Specific_Reads_Analysis/POL_PROT_RT/Quality/MutiQCResults/multiqc_data/multiqc_fastq
c.txt";
//		String RU5ReadsInfosPath
="Specific_Reads_Analysis/RU5/Quality/MutiQCResults/multiqc_data/multiqc_fastqc.txt";
//		String U3RReadsInfosPath
="Specific_Reads_Analysis/U3R/Quality/MutiQCResults/multiqc_data/multiqc_fastqc.txt";
//		String patientInfospath="Patient_info.txt";
//		String output ="results_Analysis1.csv";


// Analyse 2
		String fullReadsInfosPath
="Raw_Data_quality_Analysis_V2/MutiQCResults/multiqc_data/multiqc_fastqc.txt";
		String fullExtractReadsInfosPath
="Extract_Reads_V2/Quality/MutiQCResults/multiqc_data/multiqc_fastqc.txt";
		String ENVReadsInfosPath
="Specific_Reads_Analysis_V2/ENV/Quality/MutiQCResults/multiqc_data/multiqc_fastqc.txt";
		String ENV_U3RReadsInfosPath
="Specific_Reads_Analysis_V2/ENV_U3R/Quality/MutiQCResults/multiqc_data/multiqc_fastqc.
txt";
		String GAGReadsInfosPath
="Specific_Reads_Analysis_V2/GAG/Quality/MutiQCResults/multiqc_data/multiqc_fastqc.txt";
		String POL_INTReadsInfosPath
="Specific_Reads_Analysis_V2/POL_INT/Quality/MutiQCResults/multiqc_data/multiqc_fastqc.t
xt";
		String POL_PROT_RTReadsInfosPath
="Specific_Reads_Analysis_V2/POL_PROT_RT/Quality/MutiQCResults/multiqc_data/multiqc_fa
stqc.txt";
		String RU5ReadsInfosPath
="Specific_Reads_Analysis_V2/RU5/Quality/MutiQCResults/multiqc_data/multiqc_fastqc.txt";
		String U3RReadsInfosPath
="Specific_Reads_Analysis_V2/U3R/Quality/MutiQCResults/multiqc_data/multiqc_fastqc.txt";
		String patientInfospath="Patient_info_V2.txt";
		String output ="results_Analysis2.csv";



		try {

		BufferedWriter write = new BufferedWriter(new FileWriter(output));
		Map<String,Integer> sampleAndAllReads =
retrieveReadsPerSample(fullReadsInfosPath);
		Map<String,Integer> sampleAndExtractReads =
retrieveReadsPerSample(fullExtractReadsInfosPath);
```

```
			Map<String,Integer>			sampleAndENVReads			=
retrieveReadsPerSample(ENVReadsInfosPath);
			Map<String,Integer>			sampleAndENV_U3RReads			=
retrieveReadsPerSample(ENV_U3RReadsInfosPath);
			Map<String,Integer>			sampleAndGAGReads			=
retrieveReadsPerSample(GAGReadsInfosPath);
			Map<String,Integer>			sampleAndPOL_INTReads			=
retrieveReadsPerSample(POL_INTReadsInfosPath);
			Map<String,Integer>			sampleAndPOL_PROT_RTReads			=
retrieveReadsPerSample(POL_PROT_RTReadsInfosPath);
			Map<String,Integer>			sampleAndRU5Reads			=
retrieveReadsPerSample(RU5ReadsInfosPath);
			Map<String,Integer>			sampleAndU3RReads			=
retrieveReadsPerSample(U3RReadsInfosPath);
			Map<String,		String>		samplePacsInfos			=
retrieveSamplePacsInfos(patientInfospath);
			Map<String,		String>		sampleClusterIDInfos			=
retrieveSampleClusterIDInfos(patientInfospath);
			Set<String> AllSampleName = sampleAndAllReads.keySet();

			String line = "Full Name, Sample Name,Visit,Cell Type,ClusterID, PASC, All
Reads,Extract Reads,Env Reads,Env U3R Reads,GAG reads,Pol Int Reads,Pol Prot RT reads,RU5
Reads,U3R Reads";
			write.write(line);
			write.newLine();
			for (String string : AllSampleName)
			{
				line ="";
				String parts[] = string.split("_");
				line+=string +"," + parts[0]+","+parts[1]+","+parts[2]+",";

				line+=sampleClusterIDInfos.get(parts[0]) +",";
				line+=samplePacsInfos.get(parts[0]) +",";

				line+=sampleAndAllReads.get(string) +",";
				line+=sampleAndExtractReads.get(string) +",";
				line+=sampleAndENVReads.get(string) +",";
				line+=sampleAndENV_U3RReads.get(string) +",";
				line+=sampleAndGAGReads.get(string) +",";
				line+=sampleAndPOL_INTReads.get(string) +",";
				line+=sampleAndPOL_PROT_RTReads.get(string) +",";
				line+=sampleAndRU5Reads.get(string) +",";
				line+=sampleAndU3RReads.get(string);
				write.write(line);
				write.newLine();
				write.flush();
			}
			write.close();

			} catch (IOException e) {
				// TODO Auto-generated catch block
				e.printStackTrace();
```

```
            }

        }

        public static Map<String,Integer> retrieveReadsPerSample(String multiQCFilePath)
throws IOException
        {
            Map<String,Integer> SampleAndReads = new HashMap<String, Integer>();
            BufferedReader read = new BufferedReader(new FileReader(multiQCFilePath));
            String line ="";
            while(line!=null)
            {
                if(line.isEmpty())
                {
                    line = read.readLine();
                    continue;
                }
                if(line.startsWith("Sample"))
                {
                    line = read.readLine();
                    continue;
                }

                String[] parts = line.split("\t");
                String fullsampleName = parts[0];
                String[] fullsampleNameparts = fullsampleName.split("_");

                String shortSampleName = fullsampleNameparts[0]+"_" +
fullsampleNameparts[1]+"_" + fullsampleNameparts[2];
                Integer nbReads = Integer.parseInt(parts[4].replace(".0",""));

                SampleAndReads.put(shortSampleName, nbReads);

                line= read.readLine();
            }
            return SampleAndReads;
        }
        public static Map<String,String> retrieveSamplePacsInfos(String multiQCFilePath)
throws IOException
        {
            Map<String,String> SampleAndInfos = new HashMap<String, String>();
            BufferedReader read = new BufferedReader(new FileReader(multiQCFilePath));
            String line ="";
            while(line!=null)
            {
                if(line.isEmpty())
                {
                    line = read.readLine();
                    continue;
```

```
				}
				if(line.startsWith("sample"))
				{
					line = read.readLine();
					continue;
				}

				String[] parts = line.split("\t");
				SampleAndInfos.put(parts[0], parts[1
							]);

				line= read.readLine();
			}
			return SampleAndInfos;
		}
		public static Map<String,String> retrieveSampleClusterIDInfos(String multiQCFilePath)
throws IOException
		{
			Map<String,String> SampleAndInfos = new HashMap<String, String>();
			BufferedReader read = new BufferedReader(new FileReader(multiQCFilePath));
			String line ="";
			while(line!=null)
			{
				if(line.isEmpty())
				{
					line = read.readLine();
					continue;
				}
				if(line.startsWith("sample"))
				{
					line = read.readLine();
					continue;
				}

				String[] parts = line.split("\t");
				SampleAndInfos.put(parts[0], parts[2
							]);

				line= read.readLine();
			}
			return SampleAndInfos;
		}

}
```

Tools used for the analysis

bowtie2 v 2.2.3
fastqc v0.11.9
multiqc v1.14.dev0

Command line used for each sample analyzed

Concat all references in one Fasta file : All_references.fasta
ENV.fasta
ENV_U3R.fasta
GAG.fasta
HERW.fasta
POL_INT.fasta
POL_PROT_RT.fasta
RU5.fasta
U3.fasta
U3R.fasta
U5.fasta

### Build of bowtie reference index for each reference
### this command line was done for each fasta file of reference used during the analysis (11 times)
e.g bowtie2-build ENV.fasta ENV

### Retrieve all potential reads of interest using local alignement mode
### This command was performed for all sample analyzed (1923 time for first analysis and 2210 times for second analysis)
bowtie2 -p 18 --local -k 1 -N 1 -L 20 -i S,1,0.75 --ignore-quals --ma 4 --mp 12,12 --score-min G,-20,34 -x All_references -U SampleX_R1.fastq.gz,SampleX_R2.fastq.gz --no-unal --al Sample_Extracted_reads.fastq

### Based on reads extract we identify by a more stringent analysis reads that could be associated to each target (exemple for ENV)
bowtie2 -p 6 --end-to-end -k 1 --very-sensitive --ignore-quals -x ENV -U Sample_Extracted_reads.fastq --no-unal --al Sample_ENV_reads.fastq

#fastqc analysis to determine easlily the number of reads per files (this is done in each folder that contains all the reads extracted)
fastqc -t 12 -o Quality/ *.fastq

### multiqc analysis to retrieve all fastqc information in one report (Quality folder in same as previous command)
multiqc -o Quality/MutiQCResults/ Quality/

All the previous commands were done for each sample and for each targets

We obtain 2 folders one for the first Analysis (1923 samples) and for the second analysis (2649 samples) that contain the following folder
ENV
ENV_U3R
GAG
POL_INT
POL_PROT_RT
RU5
U3R

Then we used the following java code to retrieve all read numbers information for each samples
As input we used results of multiQC analysis results for each target analysis, raw data analysis.
We used also a text file "Patient_info.txt" that contain for each patient of the first analysis the PACS information. 'Yes' or 'No'

```
import java.io.BufferedReader;
import java.io.BufferedWriter;
import java.io.FileReader;
import java.io.FileWriter;
import java.io.IOException;
import java.util.HashMap;
import java.util.List;
import java.util.Map;
import java.util.Set;

public class FastqcAnalysisToRetreiveReadsVerily
{


	public static void main(String[] args)
	{


// ANLAYSE 1
//		String fullReadsInfosPath ="Raw_Data_quality_Analysis/MutiQCResults/multiqc_data/multiqc_fastqc.txt";
//		String fullExtractReadsInfosPath ="Extract_Reads/Quality/MutiQCResults/multiqc_data/multiqc_fastqc.txt";
//		String ENVReadsInfosPath ="Specific_Reads_Analysis/ENV/Quality/MutiQCResults/multiqc_data/multiqc_fastqc.txt";
//		String ENV_U3RReadsInfosPath ="Specific_Reads_Analysis/ENV_U3R/Quality/MutiQCResults/multiqc_data/multiqc_fastqc.txt"
;
//		String GAGReadsInfosPath ="Specific_Reads_Analysis/GAG/Quality/MutiQCResults/multiqc_data/multiqc_fastqc.txt";
//		String POL_INTReadsInfosPath ="Specific_Reads_Analysis/POL_INT/Quality/MutiQCResults/multiqc_data/multiqc_fastqc.txt";
```

```
//		String		POL_PROT_RTReadsInfosPath
="Specific_Reads_Analysis/POL_PROT_RT/Quality/MutiQCResults/multiqc_data/multiqc_fastq
c.txt";
//		String		RU5ReadsInfosPath
="Specific_Reads_Analysis/RU5/Quality/MutiQCResults/multiqc_data/multiqc_fastqc.txt";
//		String		U3RReadsInfosPath
="Specific_Reads_Analysis/U3R/Quality/MutiQCResults/multiqc_data/multiqc_fastqc.txt";
//		String patientInfospath="Patient_info.txt";
//		String output ="results_Analysis1.csv";


// Analyse 2
		String		fullReadsInfosPath
="Raw_Data_quality_Analysis_V2/MutiQCResults/multiqc_data/multiqc_fastqc.txt";
		String		fullExtractReadsInfosPath
="Extract_Reads_V2/Quality/MutiQCResults/multiqc_data/multiqc_fastqc.txt";
		String		ENVReadsInfosPath
="Specific_Reads_Analysis_V2/ENV/Quality/MutiQCResults/multiqc_data/multiqc_fastqc.txt";
		String		ENV_U3RReadsInfosPath
="Specific_Reads_Analysis_V2/ENV_U3R/Quality/MutiQCResults/multiqc_data/multiqc_fastqc.
txt";
		String		GAGReadsInfosPath
="Specific_Reads_Analysis_V2/GAG/Quality/MutiQCResults/multiqc_data/multiqc_fastqc.txt";
		String		POL_INTReadsInfosPath
="Specific_Reads_Analysis_V2/POL_INT/Quality/MutiQCResults/multiqc_data/multiqc_fastqc.t
xt";
		String		POL_PROT_RTReadsInfosPath
="Specific_Reads_Analysis_V2/POL_PROT_RT/Quality/MutiQCResults/multiqc_data/multiqc_fa
stqc.txt";
		String		RU5ReadsInfosPath
="Specific_Reads_Analysis_V2/RU5/Quality/MutiQCResults/multiqc_data/multiqc_fastqc.txt";
		String		U3RReadsInfosPath
="Specific_Reads_Analysis_V2/U3R/Quality/MutiQCResults/multiqc_data/multiqc_fastqc.txt";
		String patientInfospath="Patient_info_V2.txt";
		String output ="results_Analysis2.csv";



		try {

		BufferedWriter write = new BufferedWriter(new FileWriter(output));
		Map<String,Integer>		sampleAndAllReads		=
retrieveReadsPerSample(fullReadsInfosPath);
		Map<String,Integer>		sampleAndExtractReads		=
retrieveReadsPerSample(fullExtractReadsInfosPath);
		Map<String,Integer>		sampleAndENVReads		=
retrieveReadsPerSample(ENVReadsInfosPath);
		Map<String,Integer>		sampleAndENV_U3RReads		=
retrieveReadsPerSample(ENV_U3RReadsInfosPath);
		Map<String,Integer>		sampleAndGAGReads		=
retrieveReadsPerSample(GAGReadsInfosPath);
```

```
		Map<String,Integer>			sampleAndPOL_INTReads			=
retrieveReadsPerSample(POL_INTReadsInfosPath);
		Map<String,Integer>		sampleAndPOL_PROT_RTReads			=
retrieveReadsPerSample(POL_PROT_RTReadsInfosPath);
		Map<String,Integer>			sampleAndRU5Reads			=
retrieveReadsPerSample(RU5ReadsInfosPath);
		Map<String,Integer>			sampleAndU3RReads			=
retrieveReadsPerSample(U3RReadsInfosPath);
		Map<String,		String>		samplePacsInfos			=
retrieveSamplePacsInfos(patientInfospath);
		Map<String,		String>		sampleClusterIDInfos			=
retrieveSampleClusterIDInfos(patientInfospath);
		Set<String> AllSampleName = sampleAndAllReads.keySet();

		String line = "Full Name, Sample Name,Visit,Cell Type,ClusterID, PASC, All
Reads,Extract Reads,Env Reads,Env U3R Reads,GAG reads,Pol Int Reads,Pol Prot RT reads,RU5
Reads,U3R Reads";
		write.write(line);
		write.newLine();
		for (String string : AllSampleName)
		{
			line ="";
			String parts[] = string.split("_");
			line+=string +"," + parts[0]+","+parts[1]+","+parts[2]+",";

			line+=sampleClusterIDInfos.get(parts[0]) +",";
			line+=samplePacsInfos.get(parts[0]) +",";

			line+=sampleAndAllReads.get(string) +",";
			line+=sampleAndExtractReads.get(string) +",";
			line+=sampleAndENVReads.get(string) +",";
			line+=sampleAndENV_U3RReads.get(string) +",";
			line+=sampleAndGAGReads.get(string) +",";
			line+=sampleAndPOL_INTReads.get(string) +",";
			line+=sampleAndPOL_PROT_RTReads.get(string) +",";
			line+=sampleAndRU5Reads.get(string) +",";
			line+=sampleAndU3RReads.get(string);
			write.write(line);
			write.newLine();
			write.flush();
		}
		write.close();

		} catch (IOException e) {
			// TODO Auto-generated catch block
			e.printStackTrace();
		}



	}
```

```
	public static Map<String,Integer> retrieveReadsPerSample(String multiQCFilePath) throws IOException
	{
		Map<String,Integer> SampleAndReads = new HashMap<String, Integer>();
		BufferedReader read = new BufferedReader(new FileReader(multiQCFilePath));
		String line ="";
		while(line!=null)
		{
			if(line.isEmpty())
			{
				line = read.readLine();
				continue;
			}
			if(line.startsWith("Sample"))
			{
				line = read.readLine();
				continue;
			}

			String[] parts = line.split("\t");
			String fullsampleName = parts[0];
			String[] fullsampleNameparts = fullsampleName.split("_");

			String shortSampleName = fullsampleNameparts[0]+"_" + fullsampleNameparts[1]+"_" + fullsampleNameparts[2];
			Integer nbReads = Integer.parseInt(parts[4].replace(".0",""));

			SampleAndReads.put(shortSampleName, nbReads);

			line= read.readLine();
		}
		return SampleAndReads;
	}
	public static Map<String,String> retrieveSamplePacsInfos(String multiQCFilePath) throws IOException
	{
		Map<String,String> SampleAndInfos = new HashMap<String, String>();
		BufferedReader read = new BufferedReader(new FileReader(multiQCFilePath));
		String line ="";
		while(line!=null)
		{
			if(line.isEmpty())
			{
				line = read.readLine();
				continue;
			}
			if(line.startsWith("sample"))
			{
				line = read.readLine();
				continue;
			}
```

```
                        String[] parts = line.split("\t");
                        SampleAndInfos.put(parts[0], parts[1
                                    ]);

                        line= read.readLine();
                }
                return SampleAndInfos;
        }
        public static Map<String,String> retrieveSampleClusterIDInfos(String multiQCFilePath)
throws IOException
        {
                Map<String,String> SampleAndInfos = new HashMap<String, String>();
                BufferedReader read = new BufferedReader(new FileReader(multiQCFilePath));
                String line ="";
                while(line!=null)
                {
                        if(line.isEmpty())
                        {
                                line = read.readLine();
                                continue;
                        }
                        if(line.startsWith("sample"))
                        {
                                line = read.readLine();
                                continue;
                        }

                        String[] parts = line.split("\t");
                        SampleAndInfos.put(parts[0], parts[2
                                    ]);

                        line= read.readLine();
                }
                return SampleAndInfos;
        }

}
```
